# Multimodal Analysis of Transcriptional Regulation in the Dorsolateral Prefrontal Cortex Enhances Alzheimer’s Disease Risk Gene Discovery

**DOI:** 10.64898/2026.10.29.26366737

**Authors:** Kyurhi Kim, Qiang Liu, Aron S. Buchman, Shinya Tasaki, David A. Bennett, Yanling Wang, Philip L. De Jager, Jian Hu, Jingjing Yang, Michael P. Epstein

## Abstract

Recent transcriptome-wide association studies (TWASs) of Alzheimer’s disease (AD) dementia have identified risk genes whose effects are potentially mediated through gene expression, but most do not consider other transcriptomic regulatory modalities that may contribute to AD genetic risk. To address this gap, we profiled transcriptomic quantitative traits across six modalities––gene expression, RNA stability, isoform ratios, splice junction usage, alternative TSS and polyA usage––from dorsolateral prefrontal cortex RNA-seq data tissue (n=1,035). We profiled 396,344 transcriptomic traits, which we leveraged for quantitative trait loci (xQTL) mapping and TWAS analyses of AD. We identified 25,736 xGenes (49.6% of analyzed genes) with at least one conditionally independent xQTL in separate-modality mapping, increasing signal detection by 42% relative to studying expression alone. Cross-modality mapping reduced redundant independent xQTLs by 45% while retaining signals better aligned with biological characteristics of each modality. We colocalized cross-modality xQTLs with 55 of 75 AD GWAS risk loci, including 15 risk loci detected only with non-expression modalities. TWAS analyses identified 161 independent risk genes with 59% detected through non-expression modalities and ∼30% in multiple modalities. Follow-up proteome-wide association studies using DLPFC proteomic data provided proteomic support for 26 of these independent xTWAS risk genes. Protein–protein interaction analysis further showed that 46.6% of these independent xTWAS risk genes form an interconnected network through hub genes including *MAPT, BIN1,* and *TREM2*. Our study demonstrates that studying transcriptional modalities besides expression substantially enhances the detection of AD-associated genetic signals and elucidates their underlying mechanisms.

## Introduction

Alzheimer’s disease (AD) is a progressive neurodegenerative disorder that leads to memory loss and cognitive decline, affecting an estimated 6.7 million Americans aged 65 years and older^1^. Despite its substantial public health burden, the underlying biological mechanisms are still not fully understood. Although large-scale genome-wide association studies^2–4^ (GWASs) have identified >70 risk loci for AD, most GWAS-derived risk variants reside within non-coding regions^5^ and are hard to interpret. Studies have shown that GWAS risk loci are enriched in regulatory elements^6^, such as expression quantitative trait loci (eQTLs)^7,8^, suggesting that integrating transcriptomic and genetic data could help reveal molecular mechanisms underlying GWAS signals. Accordingly, transcriptome-wide association studies (TWAS)^9–11^ have emerged as a powerful method that combines GWAS data with reference transcriptomic and genetic data to identify risk genes that influence complex traits through genetically regulated transcriptional activity in disease-relevant tissues. TWAS operates in a two-stage framework by first using reference transcriptomic and genetic data to train a regression model of gene expression on *cis*-acting genetic variants. Then the second stage of TWAS uses the estimated effect sizes of these variants to impute genetically regulated expression (GReX) levels of the gene using GWAS data, subsequently testing for the association between imputed gene expression and disease.

Although previous TWASs of AD dementia^12–15^ have identified approximately 100 risk genes, most have focused on gene expression traits derived from RNA sequencing (RNA-seq) data and therefore may miss regulatory mechanisms captured by other transcriptomic modalities, including RNA stability, isoform ratios, splice junction usage, alternative transcription start site (TSS), and alternative polyadenylation (polyA). Prior AD studies support the functional relevance of transcriptomic modalities beyond expression; for instance, alternative splicing^16,17^ and alternative polyA^18^ have been implicated in gene expression reprogramming in AD, and RNA stability^19^ has been linked to dysregulated mRNA decay programs affecting synaptic transmission proteins. More generally, Munro^20^ showed that multimodal xQTL and TWAS analyses with Geuvadis^21^ and GTEx^7^ RNA-seq data increased xGene discovery by 66% and approximately doubled the number of significant TWAS associations across a variety of psychiatric-neurologic, cardiometabolic, blood, and immune traits. Together, these observations indicate that AD risk may be mediated through multiple layers of transcriptomic regulation not captured by expression-only analyses, motivating a systematic analysis of multiple transcriptomic modalities in disease-relevant brain tissue.

In this study, we profiled molecular quantitative traits of a total of six modalities of transcriptomic variations in dorsolateral prefrontal cortex (DLPFC) tissue and then integrated these transcriptomic data with AD GWAS data to identify variants and genes that increase disease risk through such modalities. An overview of the analytical workflow is provided in Figure 1. Using raw RNA-seq data of DLPFC postmortem samples from the Religious Orders Study and Memory and Aging Project (ROS/MAP)^22,23^ cohorts, we first utilized the computational tool Pantry^20^ to generate RNA traits, including expression, RNA stability, isoform ratios, splice junction usage, alternative TSS and polyA usage for each gene. Next, we mapped xQTLs for all six modalities using tensorQTL^24^, separately per modality (separate-modality) and jointly across all modalities (cross-modality). We performed colocalization analyses with 75 AD GWAS risk loci identified by the most recent large-scale GWAS of AD^4^ using both separate-modality and cross-modality xQTL data. We then conducted xTWAS analyses of AD using reference data of all six modalities by FUSION^25^, followed by fine mapping using the Genomic Integration Fine-mapping Tool^26^ (GIFT). Finally, to evaluate proteomic support for fine-mapped independent xTWAS risk genes, we conducted proteome-wide association studies(PWAS-O) using proteomics data^27^ of DLPFC.

**Figure 1.**
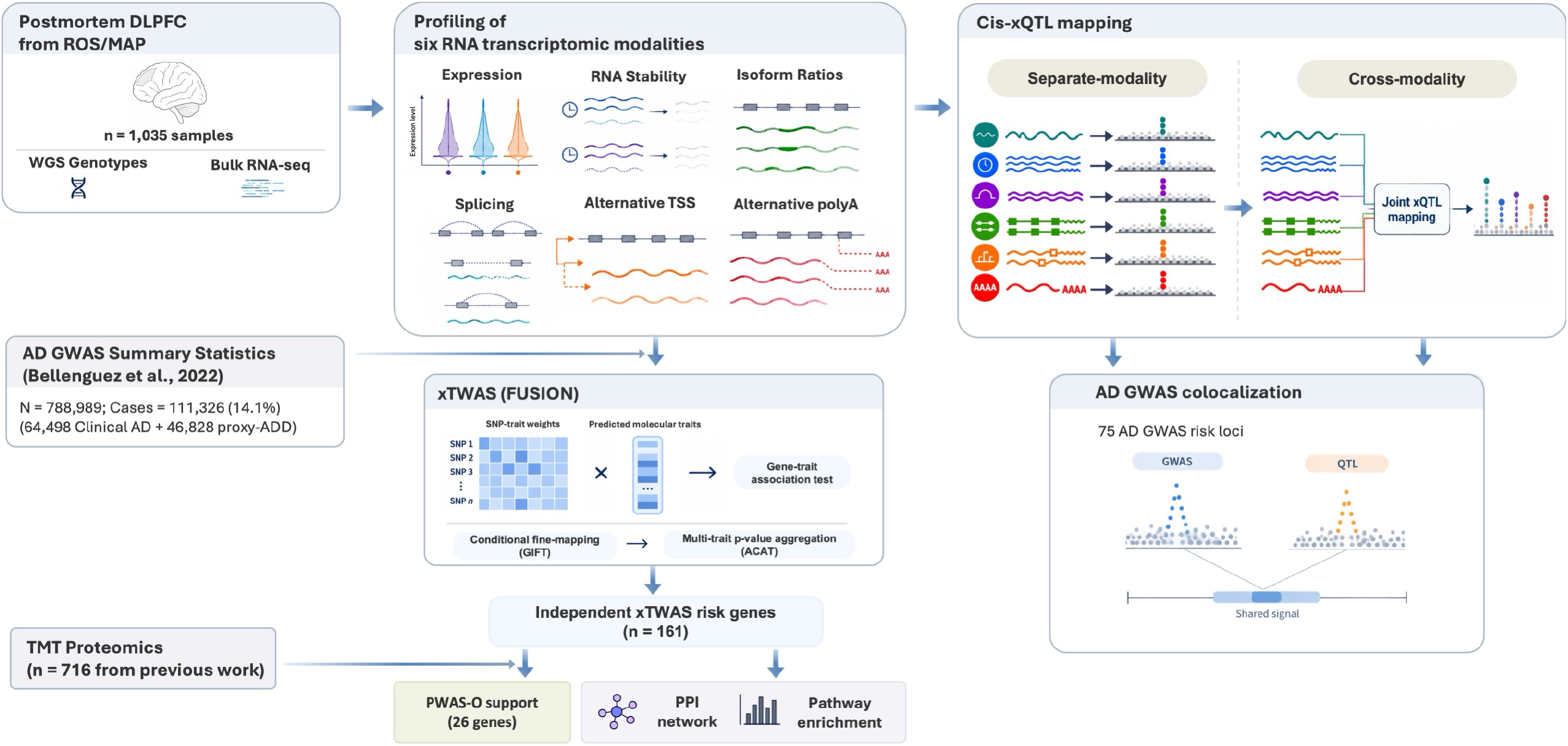
Multimodal transcriptomic analysis workflow for AD risk-gene discovery. Schematic overview of the analytical workflow. The icons were generated using ChatGPT (OpenAI, GPT-6, accessed Oct 2026), and the figure was subsequently edited and finalized by the authors using Microsoft PowerPoint.

We demonstrate that incorporating transcriptomic modalities beyond expression substantially increases discovery of conditionally independent xQTLs and identifies xTWAS risk genes for AD that would be missed by expression-only analyses. These findings highlight the value of leveraging the full regulatory information embedded in RNA-seq data to improve genetic signal discovery and better characterize the genetic architecture of AD. In the following sections, we first describe the study data and methods, followed by the xQTL and xTWAS results using the ROS/MAP DLPFC RNA-seq data^22,23^ and AD GWAS summary data^4^. We end with a discussion of the importance of studying transcriptional regulatory modalities beyond gene expression to better understand the underlying genetic etiology of complex human diseases.

## Materials and Methods

### ROS/MAP omics data

We utilized large-scale, multi-omics data from two prospective longitudinal cohort studies of aging and dementia (ROS and MAP), including genetics data profiled by whole genome sequencing (WGS), as well as bulk transcriptomics and proteomics data from postmortem DLPFC tissue^22,23,27^. Both ROS and MAP cohorts enroll participants without known dementia who agree to annual clinical evaluations and brain donation upon death; both studies were approved by the Institutional Review Board of Rush University Medical Center. All participants signed informed and repository consents, and an Anatomic Gift Act. The bulk transcriptomics data were profiled by RNA-seq, where TruSeq stranded sequencing libraries were prepared and sequenced on an Illumina NovaSeq 6000 platform with 40-50M total reads, 2×150bp paired end as previously reported^28^.

The bulk proteomics data were profiled by Tandem Mass Tag Mass Spectrometry (TMT-MS) and discussed previously^29^. Briefly, we used protein abundance traits that were scaled to sample-specific total protein abundance and log2-transformed to improve normality. Outlier samples were removed using the iterative principal component analysis (PCA) method^29^. Proteins with missing values exceeding 50% of participants were excluded, and missing values were imputed using the cohort mean for remaining participants.

To enable xQTL and xTWAS analyses, we utilized ROS/MAP genotype data from whole-genome sequencing as previously described^23^, and filtered genetic variants to common autosomal biallelic variants with minor allele frequency (MAF) ≥ 0.01 and Hardy-Weinberg equilibrium p-values <10^−5^. We analyzed a total of 1,035 RNA-seq and 716 TMT-MS samples (with corresponding WGS genotype data); all such samples were of European ancestry.

### Quantifying multimodal transcriptomic molecular traits from raw RNA-seq data

We utilized the PAN-TRanscriptomic phenotYping (Pantry) tool^20^ to profile multimodal RNA traits of transcriptional regulation in ROS/MAP subjects from their raw RNA-seq FASTQ data. Pantry integrates existing RNA-seq data analysis tools within a Snakemake^30^ workflow management system. We first aligned raw sequence reads to the GRCh38 human reference genome using STAR^31^ with corresponding Ensembl release 113 gene annotations. We then quantified RNA traits across six distinct modalities of transcriptional regulation –– gene expression, RNA stability, isoform ratios, splice junction usage, alternative TSS and polyA usage (see details in the Supplemental Methods).

To normalize the quantified RNA traits, we generated standardized trait tables for each modality using Pantry’s two-stage quantile normalization procedure. First, sample–wise quantile normalization was applied to standardize distributional effects across samples arising from factors such as highly expressed genes and library composition. For each sample, trait values were rank-ordered and replaced by the average value across samples at the corresponding rank position. Next, a rank-based inverse-normal transformation (preserving ties) was applied to each transcriptomic trait across samples to ensure suitability for downstream linear regression. Besides gene expression and RNA stability that yield a single trait per gene, the remaining four modalities can have multiple traits per gene. Multi-trait modalities were formatted into unified BED files where genomic coordinates were uniformly anchored to the TSS of the associated gene, so that the same set of *cis*-genetic variants would be considered in the xQTL and xTWAS analyses.

### Separate-modality and cross-modality xQTL analysis

We performed *cis*-xQTL mapping for each of the six transcriptomic modalities using tensorQTL^32^ via the integrated Pheast (PHEnotype Application STreamlined) pipeline embedded within Pantry. We considered genetic variants located within a ±500 kb *cis-*window around the TSS of each gene. We first mapped *cis*-xQTL separately for each modality, accounting for the confounding covariates of top 5 principal components (PCs) of the RNA trait, top 5 genotype PCs (derived from LD-pruned genotype data with no missingness and MAF ≥ 0.05), sex, education, age of death, study type (ROS or MAP), RNA Integrity Number (RIN), and batch effects. Multiple-testing correction was performed within each transcriptomic modality using the Storey q-value procedure^33^ implemented in tensorQTL, with an FDR threshold of 0.05. Then we mapped conditionally independent *cis*-xQTLs per transcriptional trait by tensorQTL through a stepwise regression algorithm (Supplemental Methods). We analyzed four modalities that yield multiple traits per gene (isoform ratios, splice junction usage, alternative TSS and polyA usage) as per-gene groups to ensure that the identified *cis*-xQTL signals were conditionally independent across all traits annotated to the same gene.

In addition to modality-specific xQTL mapping, we used Pheast to perform cross-modality mapping within each gene by jointly analyzing all RNA traits annotated to that gene across the six modalities. This approach captures xQTLs shared across correlated traits from multiple modalities, thereby reducing redundancy among modality-specific xQTLs. We also computed the top five PCs from the concatenated trait table across all six modalities and included them as covariates along with the genotype PCs and the other sample covariates described above. We then mapped conditionally independent *cis-*xQTLs for each combined trait group per gene.

### Colocalization of xQTL and GWAS summary data of AD dementia

We further performed colocalization analysis using both separate-modality and cross-modality xQTL summary data to assess whether our mapped xQTL hits colocalized with GWAS signals within 75 AD-associated risk genes^4^. We used large-scale GWAS summary data (n=788,989) of AD dementia generated by Bellenguez et al. 2022^4^ by meta-analyses with the European Alzheimer & Dementia Biobank consortium and UK Biobank dataset. This study included 64,498 clinically diagnosed AD cases, 46,828 proxy AD and related dementia (proxy-ADD), and 677,663 controls. We note that the variants in the *APOE* gene region were capped by their GWAS p-values<1e-300 in this public GWAS summary data which limited the representation of extremely significant variants in *APOE* locus, including the well-established AD risk variants *APOE E2* (*rs7412*) and *APOE E4* (*rs429358*). In this study, we were only able to consider 16 variants in the APOE region 19:44905791-44909393, including two genome-wide significant variants rs769448 (19: 44906322; GWAS p-value = 7.81E-14) and rs769452 (19: 44907853; GWAS p-value = 7.49E-18).

We performed colocalization analysis using the *coloc.susie*^34^ method via the *coloc*(v5.2.3)^35^ R package. We first implemented the Sum of Single Conditions (SuSiE) framework^36^ to map independent xQTL and GWAS signals per locus defined as ±500 kb around the GWAS lead signal. Reference linkage disequilibrium (LD r^2^) was calculated by PLINK2(v2.00a4.3)^37^ using the ROS/MAP WGS data. Second, we computed posterior probabilities for each pair of fine-mapped xQTL or GWAS signals with respect to five hypotheses: no association with the corresponding transcriptomic trait and AD (*H*_0_), association with AD only (*H*_1_), association with transcriptomic trait only (*H*_2_), association with both transcriptomic trait and AD but distinct causal variants (*H*_3_), and association with both transcriptomic trait and AD through a shared “causal” variant (*H*_4_). We used posterior probability of *H*_4_ greater than 0.75 (PP*H*_4_ ≥ 0.75) to claim colocalization of the test xQTL and GWAS signal.

### xTWAS of AD dementia with multimodal transcriptomic traits

Using the FUSION^10^ tool integrated in the Pantry/Pheast software, we performed a standard two-stage xTWAS of AD by integrating multimodal transcriptional traits generated by Pantry with the AD GWAS summary data from Bellenguez et al. 2022^18^ (see above). We first adjusted the normalized transcriptomic traits for the same covariates as described in the above *cis*-xQTL mapping analyses, and then used the resulting regression residuals for xTWAS analyses. In the first stage of the xTWAS (Supplemental Methods), FUSION trained genetic imputation models relating *cis*-SNPs within a ±500 kb window around TSS of the corresponding gene to each RNA trait. The effect sizes of *cis*-SNPs, i.e., *cis*-SNP weights, were estimated using the best performing model with the highest cross-validation (CV) *R*^2^ among the best linear unbiased predictor (BLUP), penalized regression models with elastic net (ENET), LASSO, and a single variant model with top *cis*-SNP (Top1)^10^. Only RNA traits with significant *cis*-SNP heritability (LRT p-value < 0.01) were considered tested RNA traits and evaluated for association between their genetically regulated components and AD. In the second stage, we integrated the trained *cis*-SNP weights with AD GWAS summary statistics to test associations between each genetically regulated molecular trait and AD, using reference LD derived from the ROS/MAP WGS data. For the four modalities that have multiple transcriptomic traits per gene, we first performed xTWAS per trait and subsequently applied the Aggregated Cauchy Association Test^38^ (ACAT) method to calculate a single aggregated xTWAS p-value per gene per modality. We determined significant xTWAS hits using a Bonferroni adjusted p-value < 0.05 per modality.

### Fine mapping independent xTWAS risk genes

Standard xTWAS analyses evaluate each gene independently, which often lead to inflated false positives. Such inflation could originate from the polygenicity in GWAS summary data, high LD among nearby genetic variants tagged by true causal genetic variants, overlapped predictors of *cis*-SNPs for nearby genes, and co-expresssed nearby genes^39^. To detect independent xTWAS signals, we utilized the recently developed GIFT^26^ tool to fine map independent TWAS risk genes of AD. For each significant TWAS gene, we first defined a gene window extending ±500 kb around its TSS. When the gene windows of multiple significant TWAS genes overlapped, we merged them into a single local test region and performed gene-based conditional association testing by GIFT. We then defined independent significant TWAS risk genes as those either detected by GIFT (p-value < 0.05 with Bonferroni correction for the number of genes per local test region) or those whose gene windows did not overlap with any other significant TWAS gene windows.

### Computational support of xTWAS risk genes using PWAS

To provide computational support for independent xTWAS risk genes, we compared their genomic test regions with significant omnibus PWAS (PWAS-O) findings^27^ derived from postmortem DLPFC proteomics data from the same ROS/MAP cohorts (n=716) and AD GWAS summary statistics from Bellenguez et al. 2022^4^. Briefly, PWAS p-values were obtained using analogous two-stage PWAS frameworks implemented in TIGAR^11^, PrediXcan^9^, and FUSION^10^, and were then aggregated into an omnibus PWAS p-value per protein-coding gene using ACAT^38^. In total, 4,724 protein-coding genes were tested by PWAS-O. We considered an independent xTWAS risk gene to have proteomic support if its genomic test region overlapped with that of a protein-coding gene with a PWAS-O q-value <0.05, where q-values were computed using the Benjamini–Hochberg procedure^40^ to control the false discovery rate (FDR).

### Protein-protein interaction network and enrichment analysis

We used the STRING^41^ webtool (Version 12.0) to investigate protein-protein interaction (PPI) networks underlying all identified independent significant xTWAS risk genes. Proteins encoded by the xTWAS risk genes were represented as nodes, and the edges between nodes denoted functional associations between proteins. Edge colors corresponded to different sources of interaction evidence, including co-expression, protein homology, text mining, experiments, curated databases, gene-neighborhood, gene fusion, and gene co-occurrence. A default minimum required interaction score of 0.40 was applied to generate a PPI network. We performed PPI network analysis separately for the set of independent xTWAS risk genes from each modality and for the combined independent xTWAS risk genes across all six modalities.

To characterize the biological pathways enriched among the identified TWAS risk genes, we performed pathway enrichment analysis using PathDIP 5^42^, with respect to pathway databases of PANTHER^43^, KEGG^44^, Reactome^45^, and PathBank^46^. Pathway enrichment analysis was conducted using the list of independent xTWAS risk genes identified across all six transcriptomic modalities. Statistical significance of pathway enrichment was determined by Benjamini-Hochberg FDR-adjusted q-value < 0.05.

## Results

### Separate-modality xQTL mapping

Applying Pantry to ROS/MAP DLPFC RNA-seq data (n=1,035), we quantified 396,344 RNA traits across six transcriptional modalities, spanning 51,866 genes, including 18,557 protein-coding and 25,458 long non-coding genes (Table S1). We observed correlations among RNA traits both within and across modalities, and present example correlation matrices for four representative AD risk genes (*MAPT, ACE, BIN1,* and *APOC1*), with each gene showing a distinct pattern of correlations among its RNA traits (Figure S1).

Through separate-modality xQTL mapping, we identified 80,363 significant conditionally independent signals for 25,736 (49.6%) unique genes across six modalities, including ∼18K xQTL genes (xGenes) for total gene expression, ∼9K for isoform ratio, and ∼7.7K for alternative polyA (Table S2; Figure 2A). We identified 7,593 xGenes related to modalities beyond gene expression, representing a 41.85% increase in xGene detection compared to analysis of gene expression alone. For each modality, we examined the distribution of absolute effect sizes for all conditionally independent xQTLs (Figure S2), demonstrating that the median effect sizes for xQTLs related to isoform ratio, splice junction usage, alternative TSS, and polyA usage were larger in magnitude than the median effect size for xQTLs of total gene expression. We note that this trend may be influenced by differences in statistical power across modalities due to variation in the number of traits tested per gene.

**Figure 2.**
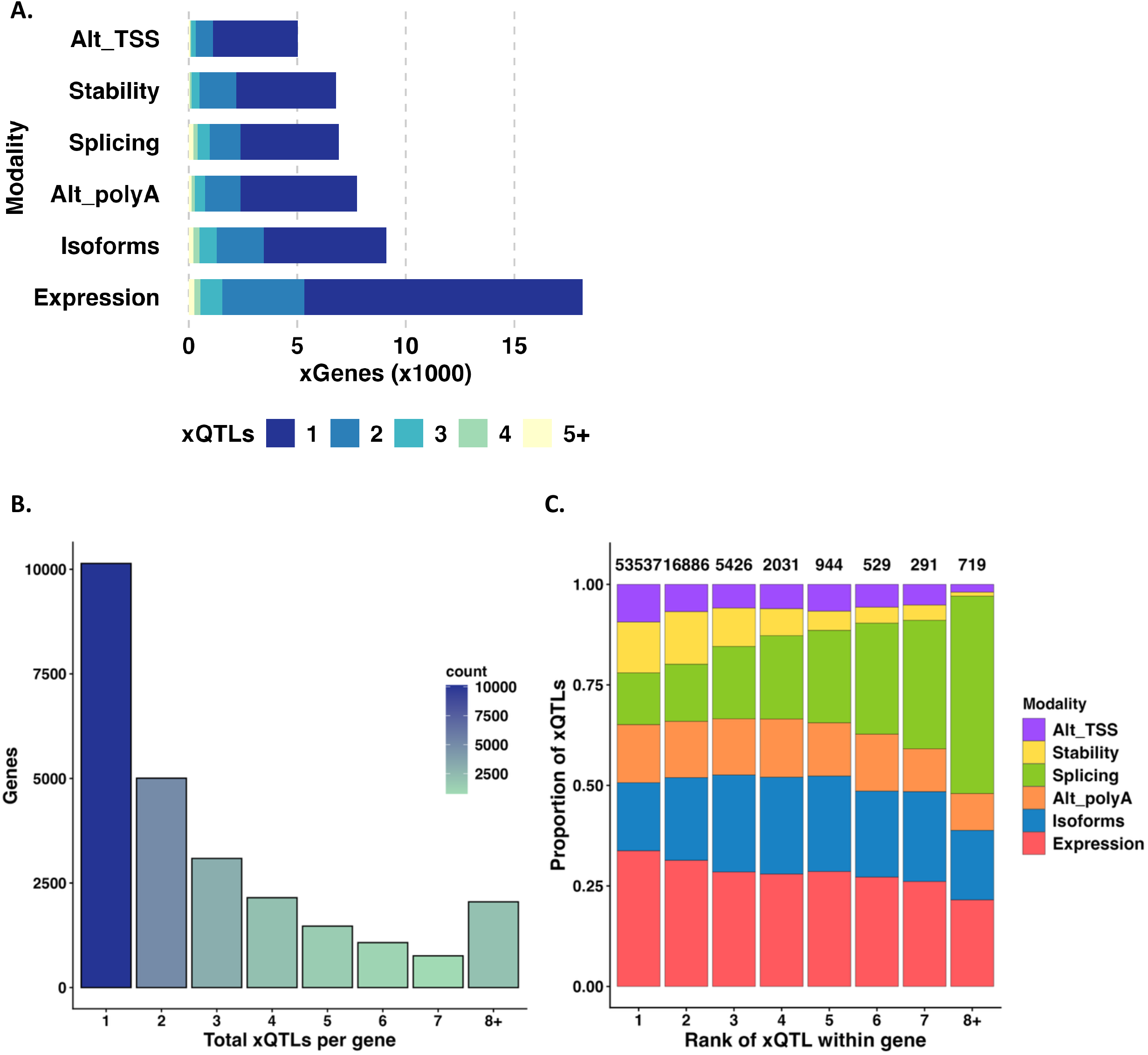
Characteristics of mapped xQTL of six transcriptional modalities by separate-modality analyses. (A) Number of unique xGenes per modality, colored by the count of independent xQTLs. (B) Histogram of the number of xGenes according to the number of their independent xQTL signals, summed over all six modalities. (C) Modality proportions of xQTLs among xQTL signals per within-xGene rank. TensorQTL assigns rank to independent QTLs for each xGene in order of their detections by stepwise regression. Rank 1 captures the top primary signal per xGene, with subsequent ranks representing conditionally independent effects of decreasing strength. The number above the bars indicate the number of independent xQTLs per rank.

We also found that 15,596 (60.6%) xGenes had 2+ conditionally independent xQTL signals (Figure 2B), with the highest proportion for the isoform ratio modality (38%) and the lowest proportion for the alternative TSS modality (22.4%) (Table 1). For example, known AD risk gene *CLU* contained two conditionally independent xQTLs for alternative polyA, one for expression, and one for alternative TSS. For each gene, tensorQTL (implemented in Pantry) assigned a rank to each conditionally independent xQTL based on their order of detection by stepwise regression, where rank 1 represents the primary signal and subsequent ranks denote weaker signals. Figure 2C shows the distribution of RNA traits across these different signal ranks. We observed that the relative proportion of xQTLs within each modality varied substantially across signal ranks. Expression QTLs constituted the largest portion of top-ranked signals, followed by isoform, alternative polyA, and splicing QTLs. Consistent with the patterns reported by Munro et al. for Geuvadis data^20^, stronger xQTLs were more likely to be associated with expression, whereas other modalities contributed more frequently to secondary and subsequent signals.

**Table 1.**
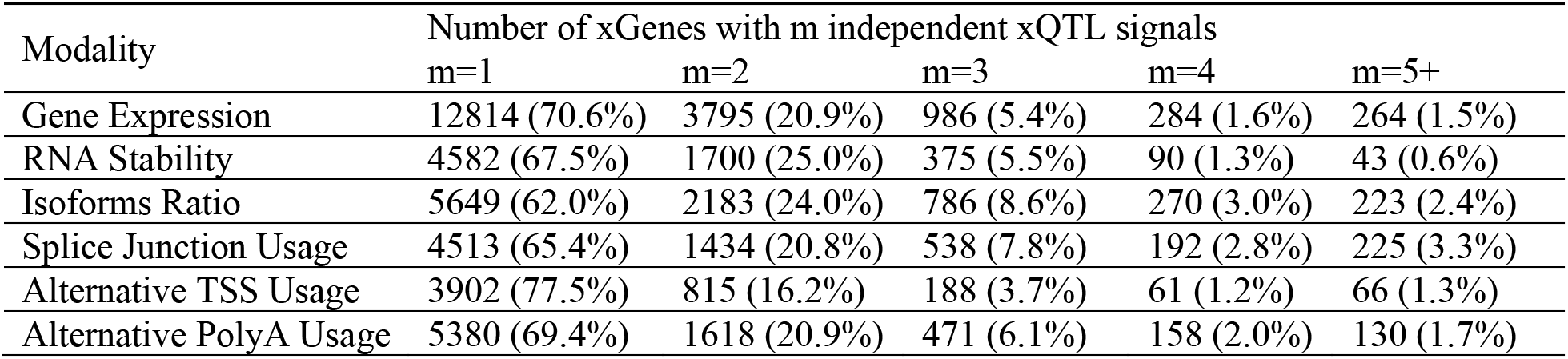
Number of xGenes with m=1, 2, 3, 4, or 5+ conditionally independent xQTL signals by separate-modality xQTL mapping. Percentages in each cell show the proportion of xGenes within that modality.

To further characterize mapped conditionally independent QTLs across the six modalities, we compared the distributions of absolute effect sizes and the distances to the corresponding TSS for primary (rank 1), secondary (rank 2), and tertiary-or-higher (tertiary+, rank ≥3) xQTLs among 3685 xGenes with at least three independent xQTL signals (Figure S3). We demonstrated that primary xQTLs were located closer to the TSS, while secondary and tertiary+ signals were positioned further away (Figure S3A). For example, the median distance to the TSS for primary expression xQTLs was 33.69 kb, significantly lower than the median 46.59 kb for secondary signals and 83.06 kb for tertiary+ signals. This trend was consistent and statistically significant across all six modalities. We also observed that, other than Alternative TSS traits, the primary xQTLs generally exhibited larger absolute effect sizes than the remaining xQTLs, with a consistent attenuation observed for subsequent secondary and tertiary+ xQTLs (Figure S3B).

Next, by examining the positional distribution of conditionally independent xQTLs per modality (Figure S4), we observed clear modality-dependent differences in the relative positions of xQTL within their xGenes. Specifically, QTLs of expression and alternative TSS were more enriched near the TSS; xQTLs of stability, isoforms, and splicing were more evenly distributed across the gene body, with modest enrichment near the TSS and gene end; while xQTLs of alternative polyA were concentrated toward the gene end. These patterns suggest that the detected signals reflect the expected regulatory mechanisms of each modality.

### Cross-modality xQTL mapping

To evaluate the impact of cross-modality xQTL mapping, we compared the number of independent xQTLs per xGene from cross-modality mapping with those obtained from separate-modality mapping (Figure 3A). Cross-modality mapping resulted in fewer total independent xQTLs per gene on average, consistent with the expectation that it removes redundant signals arising from correlated traits per gene. Despite the considerable 44.5% reduction in the total number of independent xQTLs (Figure 3B, 3C), the total number of xGenes decreased by only 7.5%. A substantial reduction of the total number of xGenes was observed within each modality, suggesting that redundant signals were removed across modalities.

**Figure 3.**
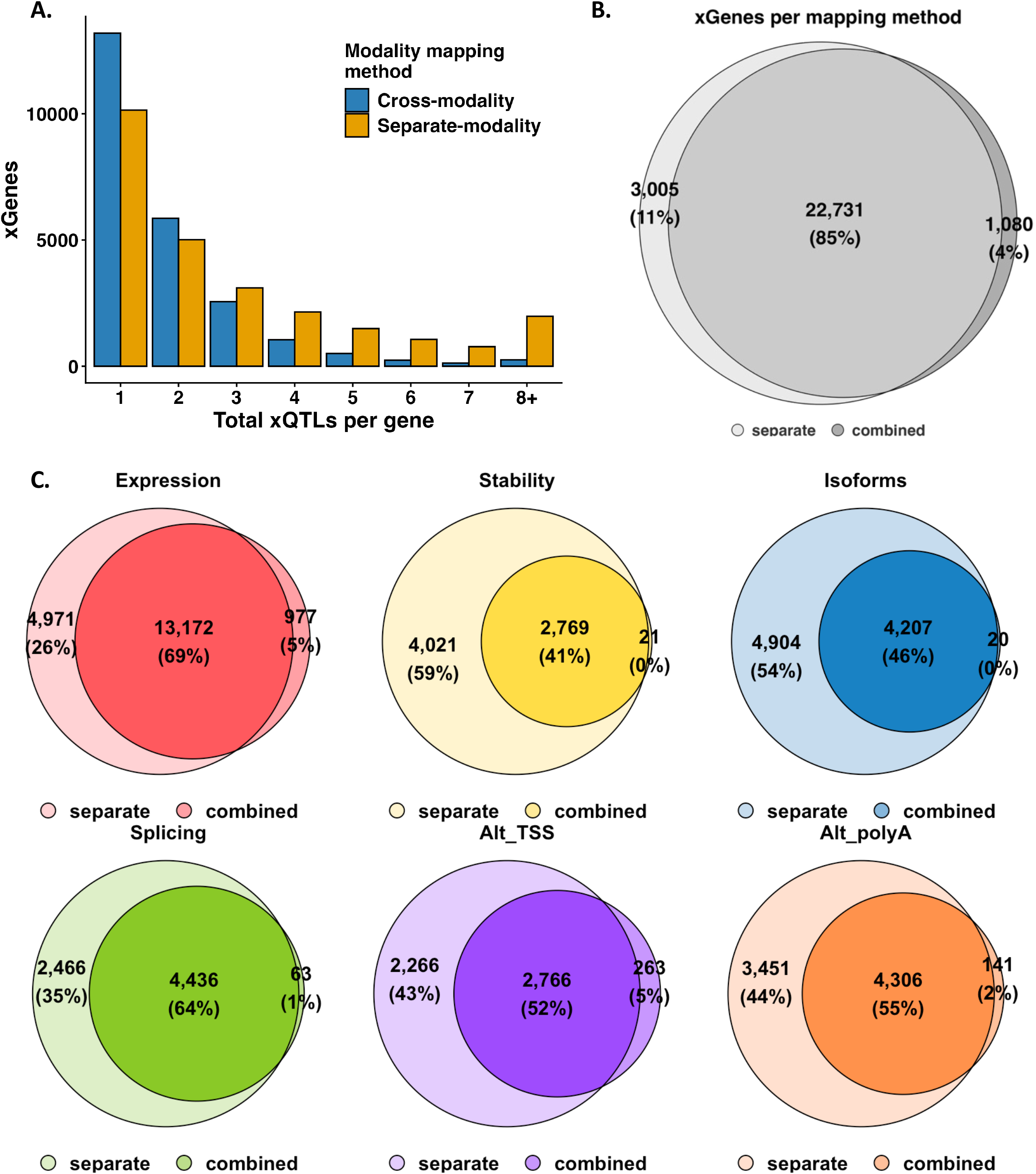
Comparing xQTL mapping results by cross-modality and separate-modality analyses. (A) Histogram of xGene numbers according to the numbers of their independent xQTLs, summed over modalities (B) Venn diagram comparing xGenes identified from separate-modality and cross-modality mapping across all six modalities (C) Venn diagram comparing xGenes identified from separate-modality and cross-modality mapping per modality. Redundant signals are removed by cross-modality analyses.

We further compared the positional distributions of xQTLs retained by cross-modality mapping, versus the positional distributions of redundant xQTLs that were removed after cross-modality mapping (Figure 4). We observed that xQTLs retained by cross-modality mapping better reflected the biological mechanisms of the corresponding transcriptional modality. For example, xQTLs related to gene expression were more clearly enriched near the TSS (gene start) in retained xQTLs versus removed xQTLs. Likewise, retained alternative polyA xQTLs were more clearly concentrated toward transcription end sites (gene end) than removed xQTLs.

**Figure 4.**
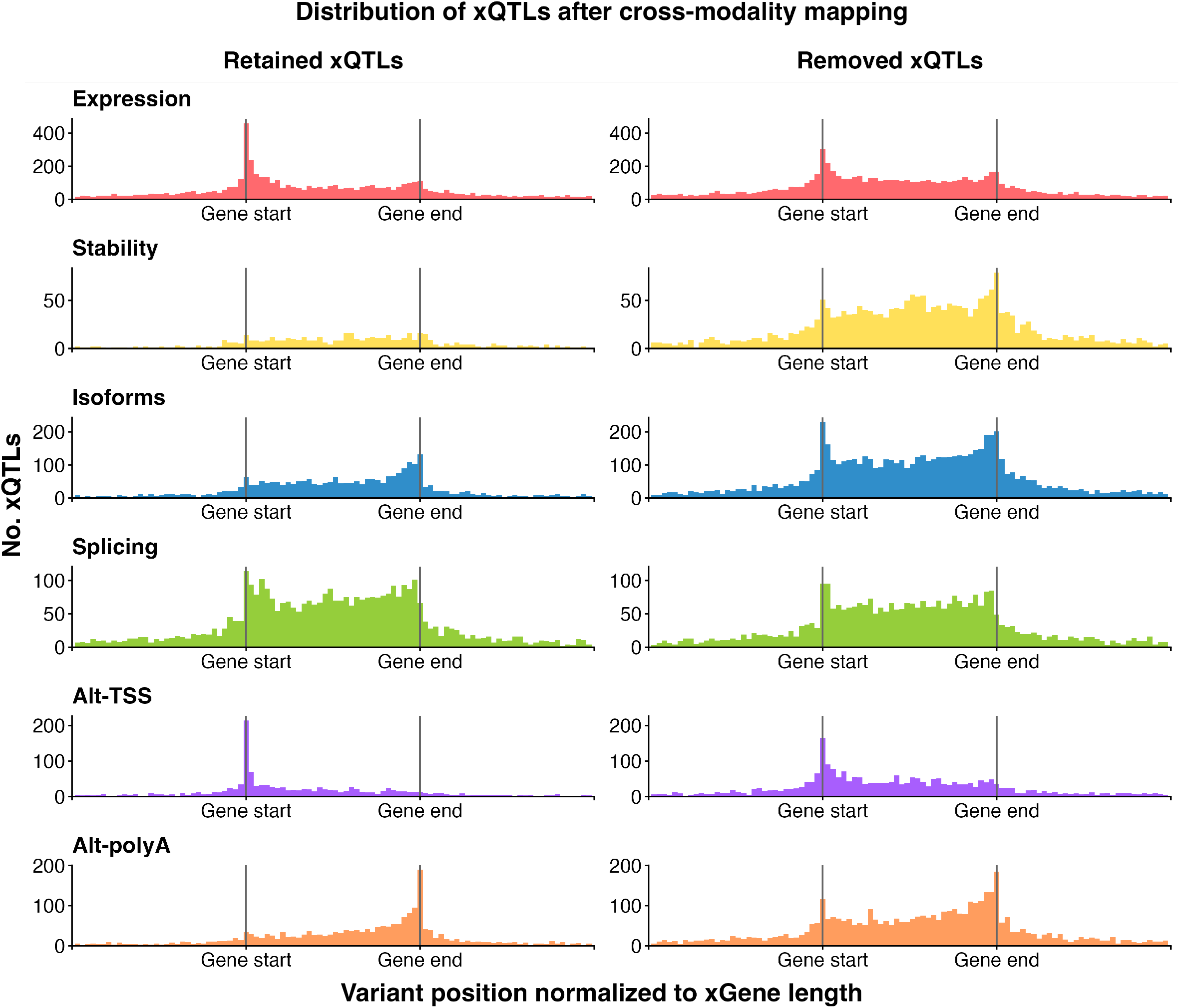
Positional distributions of xQTLs that were identified by separate-modality mapping and retained by cross-modality mapping (left column), versus the redundant ones removed (right column) by cross-modality mapping.

### Colocalizing xQTL and AD GWAS signals

To demonstrate how the xQTLs of six transcriptional modalities of DLPFC could help interpret known AD GWAS signals, we first conducted colocalization analyses leveraging separate-modality xQTL summary data and recent AD GWAS summary data^4^. Assuming a posterior probability colocalization threshold of PP*H*_4_ ≥ 0.75, we found that GWAS signals in 54 out of 75 (72%) AD GWAS risk loci^4^ colocalized with at least one xQTL signal in the six modalities, with the proportion of colocalized AD GWAS risk loci ranging from 17 (22.7%) to 35 (46.7%) in individual modalities (Table S3, Data S1). Among these 54 colocalized AD risk loci, 37 (68.5%) were colocalized in 2+ modalities (Figure S5), and 19 loci (35.2%) colocalized exclusively with non-expression modalities.

We repeated the colocalization analysis using the cross-modality xQTL summary data to assess the impact of filtering redundant signals arising from correlated RNA traits. Using the same posterior probability colocalization threshold as earlier, we identified 55 colocalized AD GWAS risk loci, of which 24 colocalized to 2+ modalities, consistent with the removal of redundant xQTLs across modalities (Figure 5). Among these 55 loci, 40 were colocalized with expression, while 15 (27.3%) were colocalized exclusively with non-expression modalities and therefore would have been missed by expression-based analyses alone (Table S3, Data S2). These findings demonstrate that non-expression modalities continue to make an important, non-redundant contribution to interpreting AD GWAS signals even after cross-modality refinement of xQTL signals.

**Figure 5.**
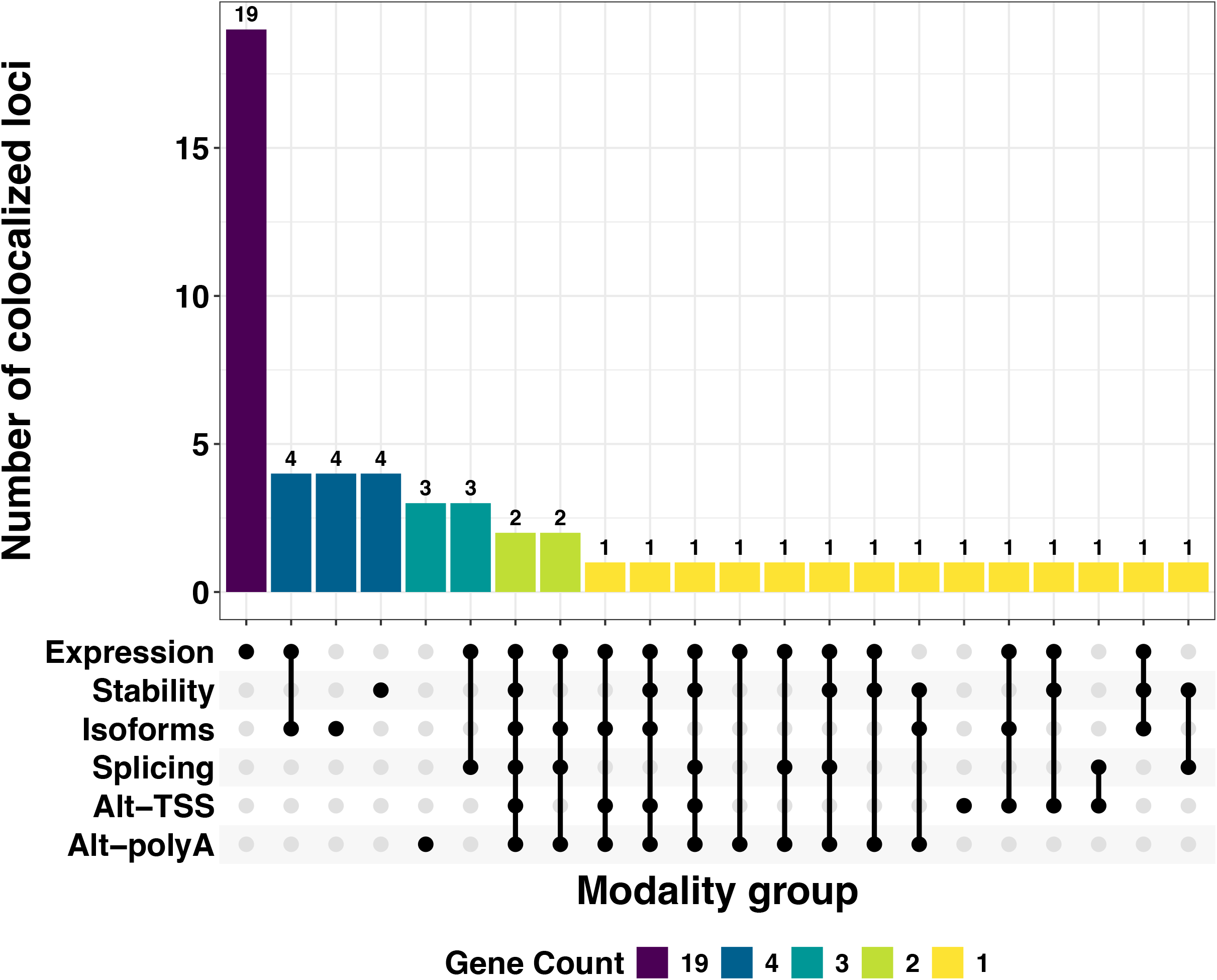
UpSet plot of AD GWAS risk loci colocalized in six modalities using the xQTL summary data from cross-modality analyses. Bar color corresponds to the number of colocalized AD GWAS risk loci. The vertical bars represent the number of colocalized loci within each specific intersections of modalities (connected dots in the bottom), as denoted by the connected dots below.

### Detection and fine mapping of xTWAS risk genes for AD

We next used FUSION (implemented in Pantry) to perform xTWAS that integrated AD GWAS summary data^4^ with the multimodal RNA-seq data that we generated from ROS/MAP. For each modality, Table S4 summarizes the number of tested RNA traits, the median cross-validation (CV) R^2^ for the tested RNA traits (i.e., estimate of RNA trait heritability due to *cis*-SNPs), the number of tested xTWAS genes, and the number of significant xTWAS risk genes. Figure S6 displays respective Manhattan plots of xTWAS results of AD per modality. Applying the Bonferroni-adjusted significance threshold per modality, we identified 214 significant xTWAS genes across all six modalities (Figure S7), including 125 genes that were associated exclusively in non-expression modalities, as well as 58 genes associated with multiple modalities.

To further fine-map independent significant xTWAS genes, we applied the GIFT tool^26^ within each local test region (see Methods). We identified 161 independent significant xTWAS genes (Figure 6, Data S3-S8), defined within each modality as those with non-overlapping test genetic regions with any other significant xTWAS genes (Table S5) and those fine-mapped by GIFT (Figure S8-S16). Of these 161 genes, 95 of them were associated exclusively with non-expression modalities, indicating that approximately 59% of detected associations would have been missed in a standard TWAS that considered only expression. Additionally, we note that 48 significant genes possessed signals in multiple modalities, suggesting a shared genetic etiology across multimodal RNA traits.

**Figure 6.**
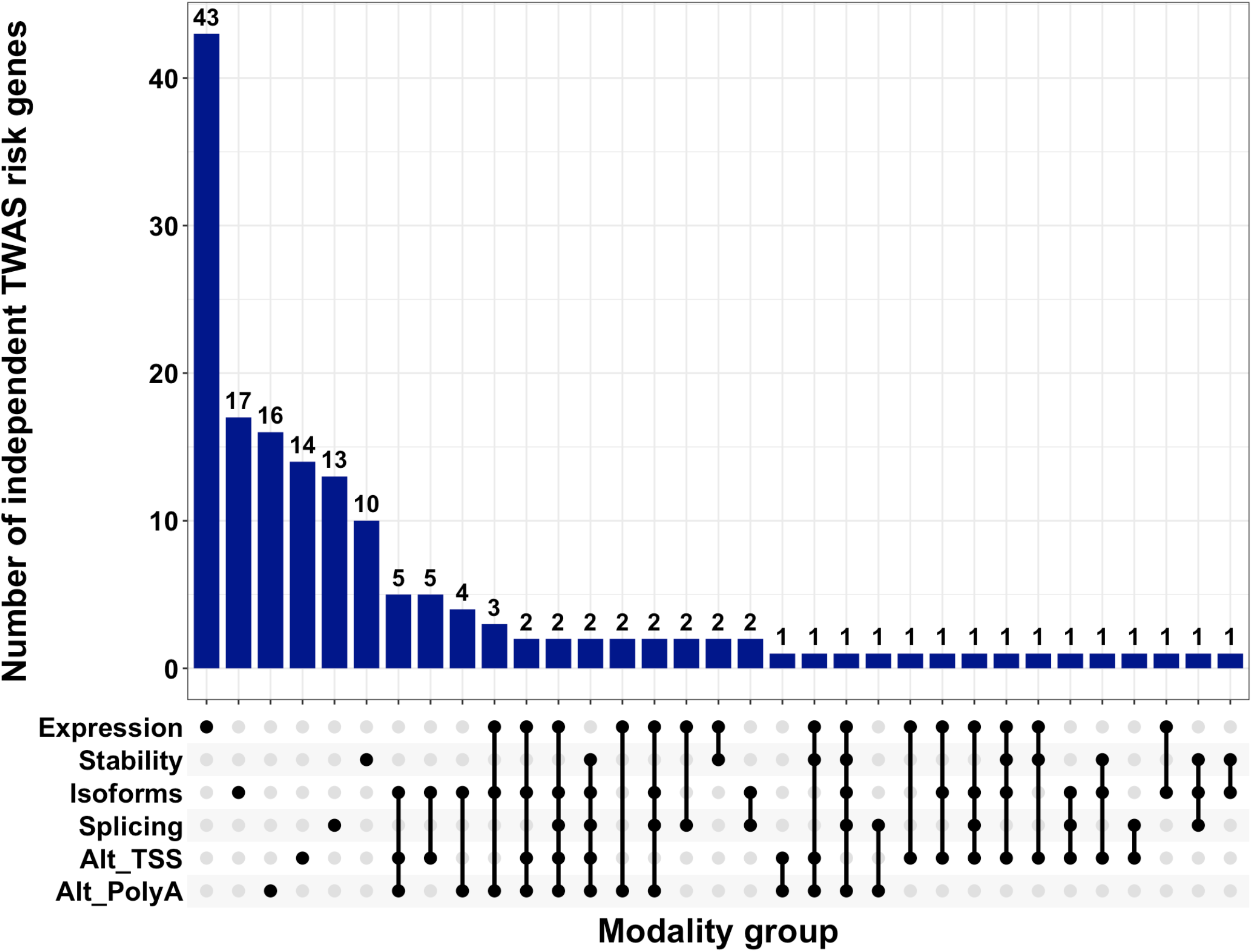
UpSet plot of independent xTWAS risk genes of AD in six transcriptional modalities. The vertical bars represent the number of independent xTWAS risk genes within each specific intersection of modalities, as denoted by the connected dots in the bottom.

### Independent xTWAS risk genes of AD supported in proteomics data

We considered independent xTWAS risk genes to have proteomic support when their test regions overlapped those of significant PWAS-O risk genes^27^ based on DLPFC proteomics data from the same ROS/MAP cohort (see Methods). Among the 161 independent xTWAS risk genes, 64 (39.8%) had genomic regions overlapping genes tested by PWAS-O and could therefore be evaluated at the protein level. Using a PWAS-O significance threshold of q < 0.05, 26 of these 64 genes (40.6%) showed proteomic support across the six modalities (Data S9, Figure S17). Of these 26 genes, 17 (65.4%) were identified exclusively through non-expression modalities, indicating that more than half would have been missed by expression-only TWAS analyses. For example, *ZKSCAN1* was identified exclusively through isoform and alternative polyA modalities. *ZKSCAN1*, a zinc-finger transcription factor, has been reported to modulate GABA type-A receptors in the brain^47^ and was identified as a master dysregulation hub gene whose coexpression networks with other brain regions are rewired in AD^48^. Another example is *APOC2*, which was identified through isoform, alternative TSS, and alternative polyA modalities. *APOC2*, which regulates lipid metabolism and cholesterol transport, is located near *APOE,* a well-known AD risk gene affecting Amyloid-β aggregation and clearance, tau pathology, and neuroinflammation^49,50^. These observations highlight the value of incorporating non-expression modalities for understanding AD-relevant genetic regulation beyond gene expression. Additionally, six of the 26 genes were supported across multiple modalities. While *ZKSCAN1* and *APOC2* were identified across multiple non-expression modalities as described above, the remaining four genes (*NDUFS2, LACTB, ICA1L,* and *ENSG00000286923*) were supported by both expression and non-expression modalities. In particular, *NDUFS2* encodes a subunit of mitochondrial complex I, and *LACTB* encodes a mitochondrial serine protease that regulates mitochondrial membrane dynamics. Mitochondrial dysfunction and oxidative stress are known to play critical roles in the onset and progression of AD^51,52^, providing a potential link between these genes and AD-related neurodegeneration. *ICA1L* is involved in vesicle trafficking and blood-brain barrier (BBB) integrity, and its downregulation has been associated with vascular leakage, white matter hyperintensities and AD^53^. *ENSG00000286923*, annotated as the long non-coding RNA *ZKSCAN1-1*, likely provides *cis*-regulatory control for the adjacent *ZKSCAN1* gene.

### PPI network and enrichment analyses of independent xTWAS risk genes of AD

Using the STRING webtool, we performed PPI network analyses using 161 independent xTWAS risk genes in all six modalities (Figure 7A), for illustrating shared biological processes. PPI edges represent predicted functional associations and were colored according to different data sources, including co-expression, experimental data, curated databases, gene fusion, co-occurrence, neighborhood, and text mining. Of the 161 genes, 107 were represented in the STRING network, of which 75 genes (46.6%) formed interconnected PPI networks (PPI enrichment p-value < 1.0 × 10^−16^). This integrated network contained clusters centered around well-established AD risk genes, including *MAPT, PICALM, APOC2, BIN1, TREM2, ABCA7, CRHR1,* and *ACE*. These findings support that multimodal transcriptional traits share biological networks, and that gene regulations affect AD genetic risk factors through multiple transcriptional mechanisms.

**Figure 7.**
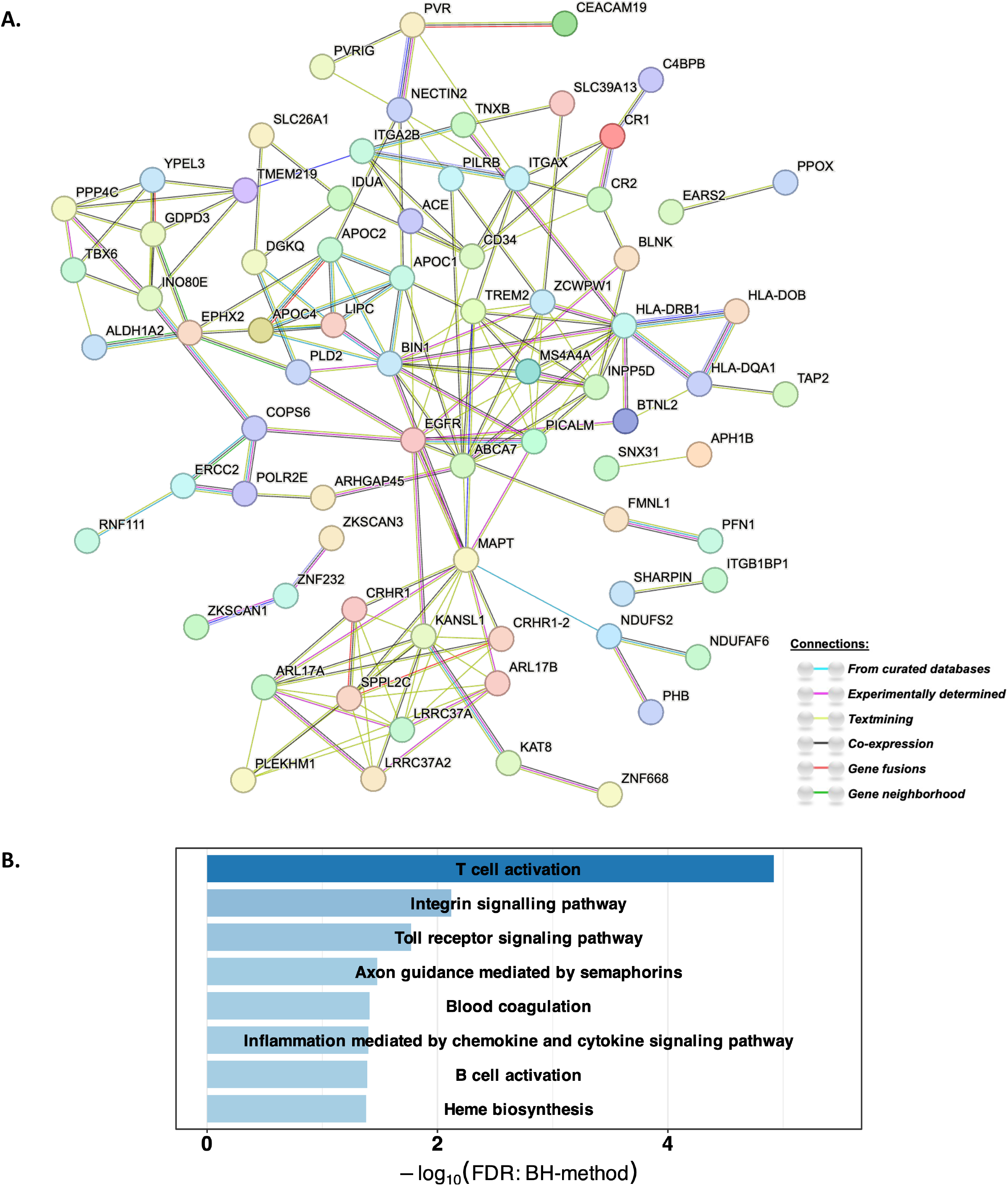
PPI networks (A) and enriched pathways (B) of 161 independent xTWAS risk genes of AD in six transcriptional modalities. (A) Edges represent PPI links, with different colors representing different sources of connection evidence. (B) Significant gene pathways enriched with independent xTWAS risk genes of AD in six modalities. The -log10(FDR) values are plotted in the x-axis.

Additionally, pathway enrichment analysis using all 161 independent xTWAS risk genes showed significant enrichment in several AD-relevant biological pathways (Figure 7B). We observed the strongest enrichment for T cell activation (FDR q-value 1.21 × 10^−5^), a pathway implicated in AD pathogenesis through its contribution to neuroinflammation^54^, with leading risk genes of *BIN1, BLNK, EGFR, INPP5D,* and *MAPT*. Particularly, *INPP5D* encodes SHIP1, a negative regulator of immune signaling that has been implicated in AD through its role in microglial inflammasome activation, autophagy dysregulation, and neuroinflammation^55^. We observed additional significant enrichment for Toll receptor signaling pathway activation (FDR q-value 0.0169), with leading risk genes *PPP4C, EGFR, NECTIN2,* and *MICA*; B cell activation (FDR q-value 0.0408) with leading risk genes *EGFR, BLNK, INPP5D, MAPT,* and *BIN1*; and inflammation mediated by chemokine and cytokine signaling (FDR q-value 0.0397), with leading risk genes *EGFR, BLNK, ITGA2B*, and *PHKG2*). Together, these results further highlight the importance of immune and neuroinflammatory processes in AD pathogenesis.

Beyond immune-related pathways, we note that we observed enrichment of several cellular signaling pathways, including integrin signaling (FDR q-value 0.0076), with leading risk genes *EGFR, ITGA2B, ITGAX,* and *PLD2*; and axon guidance mediated by semaphorins (FDR q-value 0.0334), with leading risk genes *PLD2*, *ARHGAP45*, and *MAPT*. Axon guidance has been implicated in AD pathogenesis through the regulation of hallmark proteins, including amyloid-β and tau hyperphosphorylation^56^, while integrin signaling contributes to neuroinflammation, BBB dysfunction, and Amyloid-β toxicity^57^. Additional enriched pathways included blood coagulation (FDR q-value 0.0388) with leading risk genes *C4BPB, INPP5D*, and *ITGA2B*, and heme biosynthesis (FDR q-value 0.0417), involving *EARS2, PPOX*.

We also examined PPI networks of independent xTWAS risk genes within each individual transcriptional modality (Figure S18). Across modalities, 20.7% to 52.4% of independent xTWAS risk genes formed interconnected networks and several well-established AD risk genes – including *MAPT, PICALM, CRHR1, APOC2,* and *KANSL1* – recurred as hub genes across multiple RNA regulatory modalities. In the expression, isoform ratio, and alternative TSS modalities, 30.3% (20 out of 66), 34.6% (18 out of 53), and 30% (12 out of 40) of genes formed connected PPI networks, respectively, centered on known AD risk genes such as *MAPT* and *PICALM.* In RNA stability, splice junction, and alternative polyA modalities, 52.4% (11 out of 21), 20.7% (6 out of 29), and 28.6% (12 out of 42) genes were interconnected in PPI networks, respectively, centered on genes such as *CRHR1* and *KANSL1*. These findings show that distinct RNA regulatory modalities capture partially different interaction networks while converging on a shared set of AD-related hub genes and biological pathways.

## Discussion

This study shows that multimodal RNA phenotyping of postmortem DLPFC tissue can substantially expand AD risk-gene discovery beyond expression-only approaches. Using ROS/MAP RNA-seq data, we profiled six transcriptomic modalities, performed xQTL mapping, colocalized regulatory signals with AD GWAS loci, and conducted xTWAS analyses to identify genes whose genetically regulated RNA traits were associated with AD risk. The key findings across all six transcriptomic modalities are summarized in Figure S19. Nearly 30% of xGenes (∼7,500 genes) from separate-modality mapping were detected exclusively through non-expression modalities, and cross-modality mapping refined these signals by reducing redundant xQTLs while preserving biologically interpretable modality-specific patterns, consistent with previous findings from the Pantry study^20^. Cross-modality xQTLs colocalized with 55 of 75 reported AD GWAS risk loci, including 15 loci showing colocalization exclusively with non-expression modalities, underscoring the complementary value of multimodal RNA traits for interpreting AD genetic risk.

By integrating ROS/MAP multimodal transcriptomic data with GWAS summary statistics of Alzheimer’s disease^4^, we identified 213 significant xTWAS genes across six modalities, which were subsequently fine-mapped to 161 independent xTWAS risk genes. Following xTWAS fine mapping, the relative distribution of significant genes across modalities remained highly consistent with the initial xTWAS results, supporting robust contribution of each individual modality (Figure S20). Notably, 59% of these independent xTWAS risk genes were only detected in non-expression modalities, and nearly 30% of fine-mapped independent xTWAS risk genes were detected in multiple modalities. These findings demonstrate that incorporating regulatory modalities beyond gene expression substantially improves detection of xTWAS findings for AD dementia. We further obtained proteomic support for 26 independent xTWAS risk genes using PWAS-O results derived from DLPFC proteomic data from ROS/MAP; 17 (65.4%) of which would have been missed if we had only considered gene expression in TWAS analyses. Six of these proteomically supported genes were supported by multiple transcriptional modalities, including genes involved in mitochondrial dysfunction and bioenergetic failure (*NDUFS2, LACTB*), lipoprotein metabolism and neuroinflammation (*APOC2*), and blood-brain barrier integrity (*ICA1L*).

Using the STRING tool, we found that 20%-52% of independent xTWAS risk genes formed interaction networks within individual modalities. Integrating independent xTWAS risk genes across all six modalities further expanded functional connectivity, with approximately 47% of genes forming interaction networks centered around established AD risk genes such as *MAPT, PICALM, APOC2, BIN1, TREM2, ABCA7, CRHR1* and *ACE*. Studies have previously implicated these hub genes influencing AD through diverse biological mechanisms. *MAPT* encodes the tau protein, a key neuropathological biomarker of AD pathology^58^, and *PICALM,* which encodes a clathrin assembly protein and has been implicated in AD pathogenesis through the regulation of amyloid-β (Ac) production, transport, and clearance, as well as tauopathy and synaptic dysfunction^59^. *BIN1*(Bridging Integrator 1), one of the strongest genetic risk factors for AD, has been associated with proinflammatory responses, modulation of synaptic activity, and disease-associated microglial pathway^60,61^. *TREM2* has been implicated in AD through its roles in microglial metabolism, phagocytosis, and interaction with APOE^62^. *ABCA7* dysfunction is associated with increased Aβ production, reduced Aβ clearance, impaired microglia response to inflammation, and endoplasmic reticulum stress^63^. *CRHR1*, which encodes a G-protein-coupled receptor that binds corticotropin-releasing hormone family neuropeptides, has also been implicated in AD through Aβ production and stress-related neuronal dysfunction^64^.

Consistent with the PPI network results, independent xTWAS risk genes were significantly enriched in AD-relevant immune, neuroinflammatory, and cellular signaling pathways. Several enrichments were observed primarily among genes identified through non-expression modalities. For example, integrin signalling was significantly enriched among genes identified through alternative TSS usage and isoform ratio modalities, whereas axon guidance mediated by semaphorins was significantly enriched in the alternative TSS modality (Figure S21). These pathways were not significantly enriched in the expression modality alone, highlighting that non-expression RNA traits capture complementary biological mechanisms relevant to AD. Altogether, these results demonstrate the importance of studying multimodal transcriptional traits to fully leverage the information embedded within RNA-seq data.

Nonetheless, several limitations should be considered when interpreting our findings. First, xTWAS and xQTL analyses remain inferential and cannot establish that prioritized genes or regulatory modalities causally influence AD risk; associations may reflect LD, shared *cis*-regulatory architecture, correlated nearby genes, or polygenicity in GWAS summary statistics^65–67^. To reduce false-positive inflation, we applied conservative Bonferroni correction to account for multiple testing and used GIFT to fine map xTWAS associations and identify independent xTWAS risk genes. Second, our analyses were restricted to *cis*-SNPs within a ±500 kb window around each gene TSS. Although this *cis*-window is commonly used in TWAS studies, *trans*-regulatory effects account for a substantial proportion of RNA-trait heritability^68,69^, and incorporating both *cis*- and *trans*-xQTLs into future TWAS frameworks may improve risk-gene discovery. Third, the genetically regulated components of multiple RNA traits per gene may exhibit heterogeneous genetic effect directions. Although ACAT efficiently aggregates trait-level TWAS p-values into gene-level p-value per gene, it does not incorporate effect size direction, so future methods that jointly model multiple RNA traits per gene may further improve interpretation and power.

Additional limitations relate to sample context and generalizability. The transcriptomic and proteomic analyses were conducted in bulk postmortem DLPFC tissue from European-ancestry ROS/MAP participants, which may obscure cell-type-specific regulation and limit generalizability across brain regions, disease stages, ancestries, and living-state biology. Proteomic follow-up was limited to a subset of xTWAS genes that could be evaluated using PWAS-O results based on DLPFC data from the same ROS/MAP cohort, so independent replication in diverse cohorts, additional brain regions, and cell-type-resolved datasets will be important.

In summary, this multimodal transcriptomic framework identified AD-associated genes that would likely be missed by expression-only TWAS approaches, demonstrating that AD genetic risk may act through multiple layers of transcriptional regulation. Genes supported across multiple RNA modalities may represent particularly compelling candidates because convergent regulatory signals may reflect more robust biological involvement in disease pathogenesis. These findings support integrating diverse transcriptomic traits to better capture the complexity of AD genetic regulation and motivate future multimodal analyses across additional tissues, cell types, ancestries, and disease stages.

## Acknowledgements

This work is supported by National Institutes of Health grant awards R01AG098817 (KK, MPE, JY), RF1AG071170 (KK, MPE), R01MH126449 (MPE), and R01AG089703 (JY). ROSMAP is supported by P30AG10161, P30AG72975, R01AG17917, R01AG015819, U01AG072572, and U01AG046152. ROSMAP resources can be requested at https://www.radc.rush.edu and www.synpase.org.

## Author contributions

KK led data analysis and drafted the manuscript; Q.L. and J.H. participated in data analysis and data interpretation; A.S.B., S.T., D.A.B., Y.W., and P.L.D. collected the ROS/MAP RNA-seq data and participated in data interpretation; JY and MPE conceptualized and supervised the project; all authors edited the manuscript.

## Web resources

ROS/MAP Omics Data, https://www.synapse.org/Synapse:syn3219045

Rush Alzheimer Disease Center, https://www.radc.rush.edu/

FUSION tool, http://gusevlab.org/projects/fusion/

GWAS Summary Data of AD Dementia, https://www.ebi.ac.uk/gwas/studies/GCST90027158

STRING tool, https://string-db.org/

pathDIP, https://ophid.utoronto.ca/pathDIP/

PWAS-O Summary Data of AD Dementia, https://doi.org/10.7303/syn53498818

## Data and code availability

Analysis scripts used in this study are available from https://github.com/krkim1822/AD-multimodal-transcriptomics. Cis-xQTLs, Colocalization results, xTWAS model weights, and xTWAS summary data are available from https://doi.org/10.5281/zenodo.21923545.

## Declaration of generative AI and AI-assisted technologies in the manuscript preparation process

During the preparation of this work, the authors used ChatGPT (OpenAI) to generate icons for the schematic workflow in Figure 1. All AI-assisted content was reviewed, edited, and finalized by the authors, who take full responsibility for the content of the publication.

## References

1. 2023 Alzheimer’s disease facts and figures (2023). Alzheimers Dement 19, 1598–1695. 10.1002/alz.13016.

2. Lambert, J.-C., Ibrahim-Verbaas, C.A., Harold, D., Naj, A.C., Sims, R., Bellenguez, C., Jun, G., DeStefano, A.L., Bis, J.C., Beecham, G.W., et al. (2013). Meta-analysis of 74,046 individuals identifies 11 new susceptibility loci for Alzheimer’s disease. Nat Genet 45, 1452–1458. 10.1038/ng.2802.

3. Wightman, D.P., Jansen, I.E., Savage, J.E., Shadrin, A.A., Bahrami, S., Holland, D., Rongve, A., Børte, S., Winsvold, B.S., Drange, O.K., et al. (2021). A genome-wide association study with 1,126,563 individuals identifies new risk loci for Alzheimer’s disease. Nat Genet 53, 1276–1282. 10.1038/s41588-021-00921-z.

4. Bellenguez, C., Küçükali, F., Jansen, I.E., Kleineidam, L., Moreno-Grau, S., Amin, N., Naj, A.C., Campos-Martin, R., Grenier-Boley, B., Andrade, V., et al. (2022). New insights into the genetic etiology of Alzheimer’s disease and related dementias. Nat Genet 54, 412–436. 10.1038/s41588-022-01024-z.

5. Maurano, M.T., Humbert, R., Rynes, E., Thurman, R.E., Haugen, E., Wang, H., Reynolds, A.P., Sandstrom, R., Qu, H., Brody, J., et al. (2012). Systematic localization of common disease-associated variation in regulatory DNA. Science 337, 1190–1195. 10.1126/science.1222794.

6. Li, B., and Ritchie, M.D. (2021). From GWAS to Gene: Transcriptome-Wide Association Studies and Other Methods to Functionally Understand GWAS Discoveries. Front Genet 12, 713230. 10.3389/fgene.2021.713230.

7. GTEx Consortium, Laboratory, Data Analysis &Coordinating Center (LDACC)—Analysis Working Group, Statistical Methods groups—Analysis Working Group, Enhancing GTEx (eGTEx) groups, NIH Common Fund, NIH/NCI, NIH/NHGRI, NIH/NIMH, NIH/NIDA, Biospecimen Collection Source Site—NDRI, et al. (2017). Genetic effects on gene expression across human tissues. Nature 550, 204–213. 10.1038/nature24277.

8. Nicolae, D.L., Gamazon, E., Zhang, W., Duan, S., Dolan, M.E., and Cox, N.J. (2010). Trait-Associated SNPs Are More Likely to Be eQTLs: Annotation to Enhance Discovery from GWAS. PLOS Genetics 6, e1000888. 10.1371/journal.pgen.1000888.

9. Gamazon, E.R., Wheeler, H.E., Shah, K.P., Mozaffari, S.V., Aquino-Michaels, K., Carroll, R.J., Eyler, A.E., Denny, J.C., Nicolae, D.L., Cox, N.J., et al. (2015). A gene-based association method for mapping traits using reference transcriptome data. Nat Genet 47, 1091–1098. 10.1038/ng.3367.

10. Gusev, A., Ko, A., Shi, H., Bhatia, G., Chung, W., Penninx, B.W.J.H., Jansen, R., de Geus, E.J.C., Boomsma, D.I., Wright, F.A., et al. (2016). Integrative approaches for large-scale transcriptome-wide association studies. Nat Genet 48, 245–252. 10.1038/ng.3506.

11. Nagpal, S., Meng, X., Epstein, M.P., Tsoi, L.C., Patrick, M., Gibson, G., Jager, P.L.D., Bennett, D.A., Wingo, A.P., Wingo, T.S., et al. (2019). TIGAR: An Improved Bayesian Tool for Transcriptomic Data Imputation Enhances Gene Mapping of Complex Traits. The American Journal of Human Genetics 105, 258–266. 10.1016/j.ajhg.2019.05.018.

12. Luningham, J.M., Chen, J., Tang, S., Jager, P.L.D., Bennett, D.A., Buchman, A.S., and Yang, J. (2020). Bayesian Genome-wide TWAS Method to Leverage both cis-and trans-eQTL Information through Summary Statistics. The American Journal of Human Genetics 107, 714–726. 10.1016/j.ajhg.2020.08.022.

13. Gockley, J., Montgomery, K.S., Poehlman, W.L., Liu, Y., Gerasimov, E.S., Greenwood, A.K., Wingo, A.P., Wingo, T.S., Mangravite, L.M., and Logsdon, B.A. (2020). Brain transcriptome wide association study (TWAS) implicates 8 genes across 6 loci in Alzheimer’s disease. Alzheimer’s & Dementia 16, e044839. 10.1002/alz.044839.

14. Tang, S., Buchman, A.S., Jager, P.L.D., Bennett, D.A., Epstein, M.P., and Yang, J. (2021). Novel Variance-Component TWAS method for studying complex human diseases with applications to Alzheimer’s dementia. PLOS Genetics 17, e1009482. 10.1371/journal.pgen.1009482.

15. Parrish, R.L., Buchman, A.S., Tasaki, S., Wang, Y., Avey, D., Xu, J., De Jager, P.L., Bennett, D.A., Epstein, M.P., and Yang, J. (2024). SR-TWAS: leveraging multiple reference panels to improve transcriptome-wide association study power by ensemble machine learning. Nat Commun 15, 6646. 10.1038/s41467-024-50983-w.

16. Biamonti, G., Amato, A., Belloni, E., Di Matteo, A., Infantino, L., Pradella, D., and Ghigna, C. (2021). Alternative splicing in Alzheimer’s disease. Aging Clin Exp Res 33, 747–758. 10.1007/s40520-019-01360-x.

17. Lu, Y., Tan, L., Xie, J., Cheng, L., and Wang, X. (2022). Distinct microglia alternative splicing in Alzheimer’s disease. Aging (Albany NY) 14, 6554–6566. 10.18632/aging.204223.

18. Patel, R., Brophy, C., Hickling, M., Neve, J., and Furger, A. (2019). Alternative cleavage and polyadenylation of genes associated with protein turnover and mitochondrial function are deregulated in Parkinson’s, Alzheimer’s and ALS disease. BMC Med Genomics 12, 60. 10.1186/s12920-019-0509-4.

19. Alkallas, R., Fish, L., Goodarzi, H., and Najafabadi, H.S. (2017). Inference of RNA decay rate from transcriptional profiling highlights the regulatory programs of Alzheimer’s disease. Nat Commun 8, 909. 10.1038/s41467-017-00867-z.

20. Munro, D., Ehsan, N., Esmaeili-Fard, S.M., Gusev, A., Palmer, A.A., and Mohammadi, P. (2024). Multimodal analysis of RNA sequencing data powers discovery of complex trait genetics. Nat Commun 15, 10387. 10.1038/s41467-024-54840-8.

21. Lappalainen, T., Sammeth, M., Friedländer, M.R., ‘t Hoen, P.A.C., Monlong, J., Rivas, M.A., Gonzàlez-Porta, M., Kurbatova, N., Griebel, T., Ferreira, P.G., et al. (2013). Transcriptome and genome sequencing uncovers functional variation in humans. Nature 501, 506–511. 10.1038/nature12531.

22. Bennett, D.A., Buchman, A.S., Boyle, P.A., Barnes, L.L., Wilson, R.S., and Schneider, J.A. (2018). Religious Orders Study and Rush Memory and Aging Project. J Alzheimers Dis 64, S161–S189. 10.3233/JAD-179939.

23. De Jager, P.L., Ma, Y., McCabe, C., Xu, J., Vardarajan, B.N., Felsky, D., Klein, H.-U., White, C.C., Peters, M.A., Lodgson, B., et al. (2018). A multi-omic atlas of the human frontal cortex for aging and Alzheimer’s disease research. Sci Data 5, 180142. 10.1038/sdata.2018.142.

24. Taylor-Weiner, A., Aguet, F., Haradhvala, N.J., Gosai, S., Anand, S., Kim, J., Ardlie, K., Van Allen, E.M., and Getz, G. (2019). Scaling computational genomics to millions of individuals with GPUs. Genome Biol 20, 228. 10.1186/s13059-019-1836-7.

25. Gusev, A., Ko, A., Shi, H., Bhatia, G., Chung, W., Penninx, B.W.J.H., Jansen, R., de Geus, E.J.C., Boomsma, D.I., Wright, F.A., et al. (2016). Integrative approaches for large-scale transcriptome-wide association studies. Nat Genet 48, 245–252. 10.1038/ng.3506.

26. Liu, L., Yan, R., Guo, P., Ji, J., Gong, W., Xue, F., Yuan, Z., and Zhou, X. (2024). Conditional transcriptome-wide association study for fine-mapping candidate causal genes. Nat Genet 56, 348–356. 10.1038/s41588-023-01645-y.

27. Hu, T., Liu, Q., Dai, Q., Buchman, A.S., Bennett, D.A., Tasaki, S., Wang, Y., Seyfried, N.T., De Jager, P.L., Epstein, M.P., et al. (2025). Proteome-wide association studies using summary pQTL data of brain, CSF, and plasma identify 30 risk genes of Alzheimer’s disease dementia. Alz Res Therapy 17, 135. 10.1186/s13195-025-01774-y.

28. Iturria-Medina, Y., Adewale, Q., Khan, A.F., Ducharme, S., Rosa-Neto, P., O’Donnell, K., Petyuk, V.A., Gauthier, S., De Jager, P.L., Breitner, J., et al. (2022). Unified epigenomic, transcriptomic, proteomic, and metabolomic taxonomy of Alzheimer’s disease progression and heterogeneity. Sci Adv 8, eabo6764. 10.1126/sciadv.abo6764.

29. Wingo, A.P., Fan, W., Duong, D.M., Gerasimov, E.S., Dammer, E.B., Liu, Y., Harerimana, N.V., White, B., Thambisetty, M., Troncoso, J.C., et al. (2020). Shared proteomic effects of cerebral atherosclerosis and Alzheimer’s disease on the human brain. Nat Neurosci 23, 696–700. 10.1038/s41593-020-0635-5.

30. Mölder, F., Jablonski, K.P., Letcher, B., Hall, M.B., van Dyken, P.C., Tomkins-Tinch, C.H., Sochat, V., Forster, J., Vieira, F.G., Meesters, C., et al. (2021). Sustainable data analysis with Snakemake. F1000Res 10, 33. 10.12688/f1000research.29032.3.

31. Dobin, A., Davis, C.A., Schlesinger, F., Drenkow, J., Zaleski, C., Jha, S., Batut, P., Chaisson, M., and Gingeras, T.R. (2013). STAR: ultrafast universal RNA-seq aligner. Bioinformatics 29, 15–21. 10.1093/bioinformatics/bts635.

32. Taylor-Weiner, A., Aguet, F., Haradhvala, N.J., Gosai, S., Anand, S., Kim, J., Ardlie, K., Van Allen, E.M., and Getz, G. (2019). Scaling computational genomics to millions of individuals with GPUs. Genome Biol 20, 228. 10.1186/s13059-019-1836-7.

33. Storey, J.D., and Tibshirani, R. (2003). Statistical significance for genomewide studies. Proc Natl Acad Sci U S A 100, 9440–9445. 10.1073/pnas.1530509100.

34. Wallace, C. (2021). A more accurate method for colocalisation analysis allowing for multiple causal variants. PLOS Genetics 17, e1009440. 10.1371/journal.pgen.1009440.

35. Giambartolomei, C., Vukcevic, D., Schadt, E.E., Franke, L., Hingorani, A.D., Wallace, C., and Plagnol, V. (2014). Bayesian Test for Colocalisation between Pairs of Genetic Association Studies Using Summary Statistics. PLoS Genet 10, e1004383. 10.1371/journal.pgen.1004383.

36. Wang, G., Sarkar, A., Carbonetto, P., and Stephens, M. (2020). A simple new approach to variable selection in regression, with application to genetic fine mapping. Journal of the Royal Statistical Society: Series B (Statistical Methodology) 82, 1273–1300. 10.1111/rssb.12388.

37. Chang, C.C., Chow, C.C., Tellier, L.C., Vattikuti, S., Purcell, S.M., and Lee, J.J. (2015). Second-generation PLINK: rising to the challenge of larger and richer datasets. Gigascience 4, s13742-015-0047–0048. 10.1186/s13742-015-0047-8.

38. Liu, Y., Chen, S., Li, Z., Morrison, A.C., Boerwinkle, E., and Lin, X. (2019). ACAT: A Fast and Powerful p Value Combination Method for Rare-Variant Analysis in Sequencing Studies. The American Journal of Human Genetics 104, 410–421. 10.1016/j.ajhg.2019.01.002.

39. Liang, Y., Nyasimi, F., and Im, H.K. (2024). Pervasive polygenicity of complex traits inflates false positive rates in transcriptome-wide association studies. bioRxiv, 2023.10.17.562831. 10.1101/2023.10.17.562831.

40. Benjamini, Y., and Hochberg, Y. (1995). Controlling the False Discovery Rate: A Practical and Powerful Approach to Multiple Testing. Journal of the Royal Statistical Society. Series B (Methodological) 57, 289–300.

41. Szklarczyk, D., Kirsch, R., Koutrouli, M., Nastou, K., Mehryary, F., Hachilif, R., Gable, A.L., Fang, T., Doncheva, N.T., Pyysalo, S., et al. (2023). The STRING database in 2023: protein-protein association networks and functional enrichment analyses for any sequenced genome of interest. Nucleic Acids Res 51, D638–D646. 10.1093/nar/gkac1000.

42. Pastrello, C., Kotlyar, M., Abovsky, M., Lu, R., and Jurisica, I. (2024). PathDIP 5: improving coverage and making enrichment analysis more biologically meaningful. Nucleic Acids Res 52, D663–D671. 10.1093/nar/gkad1027.

43. Thomas, P.D., Ebert, D., Muruganujan, A., Mushayahama, T., Albou, L.-P., and Mi, H. (2022). PANTHER: Making genome-scale phylogenetics accessible to all. Protein Sci 31, 8–22. 10.1002/pro.4218.

44. Kanehisa, M., Furumichi, M., Sato, Y., Kawashima, M., and Ishiguro-Watanabe, M. (2023). KEGG for taxonomy-based analysis of pathways and genomes. Nucleic Acids Res 51, D587–D592. 10.1093/nar/gkac963.

45. Milacic, M., Beavers, D., Conley, P., Gong, C., Gillespie, M., Griss, J., Haw, R., Jassal, B., Matthews, L., May, B., et al. (2024). The Reactome Pathway Knowledgebase 2024. Nucleic Acids Res 52, D672–D678. 10.1093/nar/gkad1025.

46. Wishart, D.S., Li, C., Marcu, A., Badran, H., Pon, A., Budinski, Z., Patron, J., Lipton, D., Cao, X., Oler, E., et al. (2020). PathBank: a comprehensive pathway database for model organisms. Nucleic Acids Res 48, D470–D478. 10.1093/nar/gkz861.

47. Mulligan, M.K., Wang, X., Adler, A.L., Mozhui, K., Lu, L., and Williams, R.W. (2012). Complex Control of GABA(A) Receptor Subunit mRNA Expression: Variation, Covariation, and Genetic Regulation. PLOS ONE 7, e34586. 10.1371/journal.pone.0034586.

48. Mitra, S., Bp, K., C R, S., Saikumar, N.V., Philip, P., and Narayanan, M. (2024). Alzheimer’s disease rewires gene coexpression networks coupling different brain regions. npj Syst Biol Appl 10, 50. 10.1038/s41540-024-00376-y.

49. Shao, Y., Shaw, M., Todd, K., Khrestian, M., D’Aleo, G., Barnard, P.J., Zahratka, J., Pillai, J., Yu, C.-E., Keene, C.D., et al. (2018). DNA methylation of TOMM40-APOE-APOC2 in Alzheimer’s disease. J Hum Genet 63, 459–471. 10.1038/s10038-017-0393-8.

50. Yamazaki, Y., Zhao, N., Caulfield, T.R., Liu, C.-C., and Bu, G. (2019). Apolipoprotein E and Alzheimer disease: pathobiology and targeting strategies. Nat Rev Neurol 15, 501–518. 10.1038/s41582-019-0228-7.

51. Misrani, A., Tabassum, S., and Yang, L. (2021). Mitochondrial Dysfunction and Oxidative Stress in Alzheimer’s Disease. Front Aging Neurosci 13, 617588. 10.3389/fnagi.2021.617588.

52. Wang, S., Liao, Z., Zhang, Q., Han, X., Liu, C., and Wang, J. (2024). Mitochondrial dysfunction in Alzheimer’s disease: a key frontier for future targeted therapies. Front Immunol 15, 1484373. 10.3389/fimmu.2024.1484373.

53. Chaar, D.L., Li, Z., Shang, L., Ratliff, S.M., Mosley, T.H., Kardia, S.L.R., Zhao, W., Zhou, X., and Smith, J.A. (2025). Multi-Ancestry Transcriptome-Wide Association Studies of Cognitive Function, White Matter Hyperintensity, and Alzheimer’s Disease. International Journal of Molecular Sciences 26, 2443. 10.3390/ijms26062443.

54. Town, T., Tan, J., Flavell, R.A., and Mullan, M. (2005). T-cells in alzheimer’s disease. Neuromol Med 7, 255–264. 10.1385/NMM:7:3:255.

55. Chou, V., Pearse, R.V., Aylward, A.J., Ashour, N., Taga, M., Terzioglu, G., Fujita, M., Fancher, S.B., Sigalov, A., Benoit, C.R., et al. (2023). INPP5D regulates inflammasome activation in human microglia. Nat Commun 14, 7552. 10.1038/s41467-023-42819-w.

56. Zhang, L., Qi, Z., Li, J., Li, M., Du, X., Wang, S., Zhou, G., Xu, B., Liu, W., Xi, S., et al. (2021). Roles and Mechanisms of Axon-Guidance Molecules in Alzheimer’s Disease. Mol Neurobiol 58, 3290–3307. 10.1007/s12035-021-02311-2.

57. Biose, I.J., Ismael, S., Ouvrier, B., White, A.L., and Bix, G.J. (2023). The Potential Role of Integrin Signaling in Memory and Cognitive Impairment. Biomolecules 13, 108. 10.3390/biom13010108.

58. Allen, M., Kachadoorian, M., Quicksall, Z., Zou, F., Chai, H.S., Younkin, C., Crook, J.E., Pankratz, V.S., Carrasquillo, M.M., Krishnan, S., et al. (2014). Association of MAPT haplotypes with Alzheimer’s disease risk and MAPT brain gene expression levels. Alz Res Therapy 6, 39. 10.1186/alzrt268.

59. Xu, W., Tan, L., and Yu, J.-T. (2015). The Role of PICALM in Alzheimer’s Disease. Mol Neurobiol 52, 399–413. 10.1007/s12035-014-8878-3.

60. Sudwarts, A., Ramesha, S., Gao, T., Ponnusamy, M., Wang, S., Hansen, M., Kozlova, A., Bitarafan, S., Kumar, P., Beaulieu-Abdelahad, D., et al. (2022). BIN1 is a key regulator of proinflammatory and neurodegeneration-related activation in microglia. Mol Neurodegener 17, 33. 10.1186/s13024-022-00535-x.

61. Saha, O., Melo de Farias, A.R., Pelletier, A., Siedlecki-Wullich, D., Landeira, B.S., Gadaut, J., Carrier, A., Vreulx, A.-C., Guyot, K., Shen, Y., et al. (2024). The Alzheimer’s disease risk gene BIN1 regulates activity-dependent gene expression in human-induced glutamatergic neurons. Mol Psychiatry 29, 2634–2646. 10.1038/s41380-024-02502-y.

62. Wolfe, C.M., Fitz, N.F., Nam, K.N., Lefterov, I., and Koldamova, R. (2018). The Role of APOE and TREM2 in Alzheimerʹs Disease—Current Understanding and Perspectives. Int J Mol Sci 20, 81. 10.3390/ijms20010081.

63. Stepler, K.E., Gillyard, T.R., Reed, C.B., Avery, T.M., Davis, J.S., and Robinson, R.A.S. (2022). ABCA7, a Genetic Risk Factor Associated with Alzheimer’s Disease Risk in African Americans. J Alzheimers Dis 86, 5–19. 10.3233/JAD-215306.

64. Futch, H.S., Croft, C.L., Truong, V.Q., Krause, E.G., and Golde, T.E. (2017). Targeting psychologic stress signaling pathways in Alzheimer’s disease. Mol Neurodegener 12, 49. 10.1186/s13024-017-0190-z.

65. Sun, Y., Zhu, J., Zhou, D., Canchi, S., Wu, C., Cox, N.J., Rissman, R.A., Gamazon, E.R., and Wu, L. (2021). A transcriptome-wide association study of Alzheimer’s disease using prediction models of relevant tissues identifies novel candidate susceptibility genes. Genome Med 13, 141. 10.1186/s13073-021-00959-y.

66. Liu, N., Xu, J., Liu, H., Zhang, S., Li, M., Zhou, Y., Qin, W., Li, M.J., Yu, C., and Initiative, for the A. disease N. (2021). Hippocampal transcriptome-wide association study and neurobiological pathway analysis for Alzheimer’s disease. PLOS Genetics 17, e1009363. 10.1371/journal.pgen.1009363.

67. Wainberg, M., Sinnott-Armstrong, N., Mancuso, N., Barbeira, A.N., Knowles, D.A., Golan, D., Ermel, R., Ruusalepp, A., Quertermous, T., Hao, K., et al. (2019). Opportunities and challenges for transcriptome-wide association studies. Nat Genet 51, 592–599. 10.1038/s41588-019-0385-z.

68. Guo, S., and Yang, J. (2024). Bayesian genome-wide TWAS with reference transcriptomic data of brain and blood tissues identified 141 risk genes for Alzheimer’s disease dementia. Alz Res Therapy 16, 120. 10.1186/s13195-024-01488-7.

69. Võsa, U., Claringbould, A., Westra, H.-J., Bonder, M.J., Deelen, P., Zeng, B., Kirsten, H., Saha, A., Kreuzhuber, R., Yazar, S., et al. (2021). Large-scale cis- and trans-eQTL analyses identify thousands of genetic loci and polygenic scores that regulate blood gene expression. Nat Genet 53, 1300–1310. 10.1038/s41588-021-00913-z.

