## Supplemental_Figure_Table for "Multimodal Analysis of Transcriptional Regulation in the Dorsolateral Prefrontal Cortex Enhances Alzheimer’s Disease Risk Gene Discovery"

### Supplementary Figures

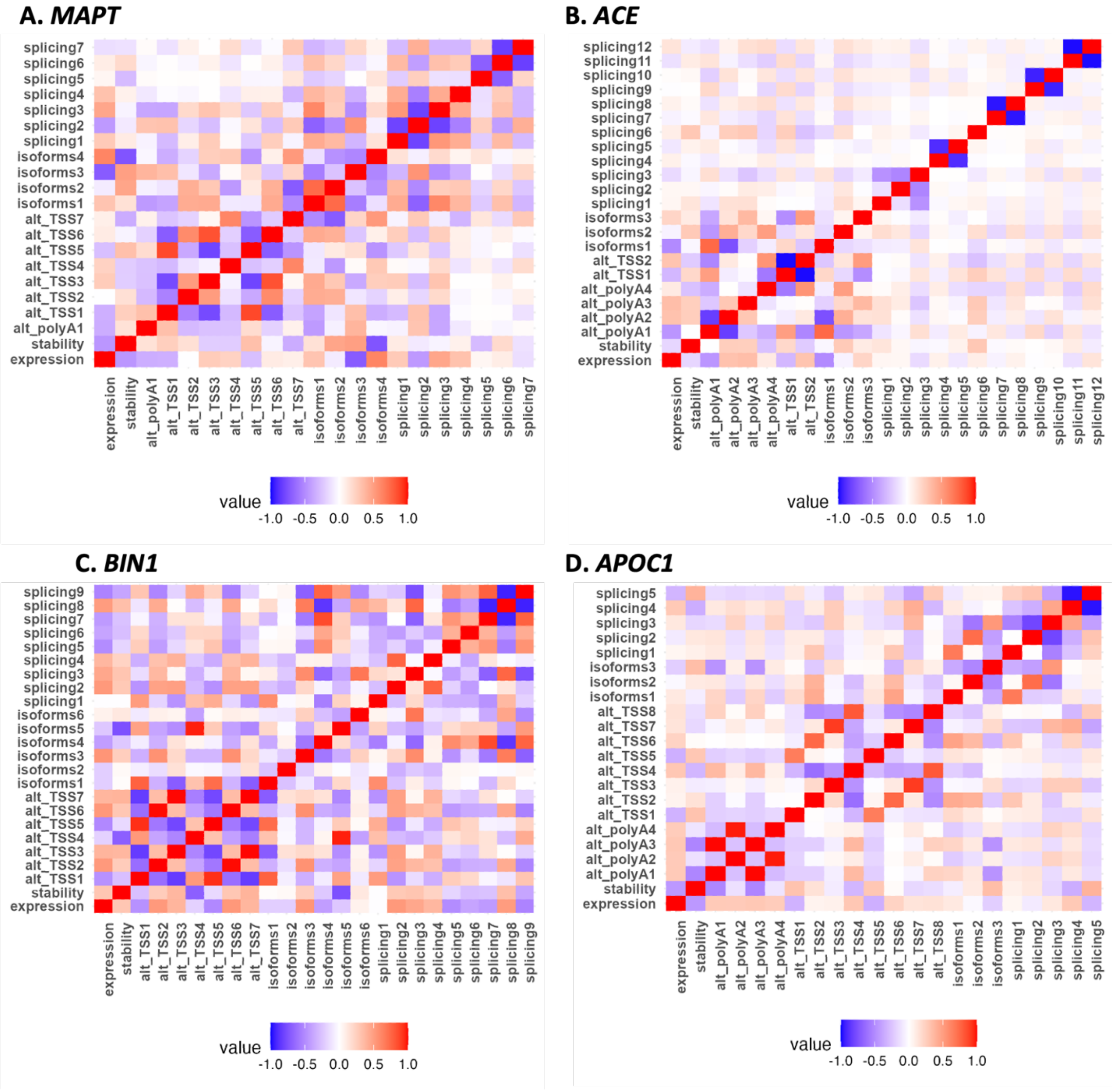


**Figure S1. Correlation heatmaps among profiled multimodal RNA traits for four example AD risk genes: *MAPT, ACE, BIN1, APOC1*.**

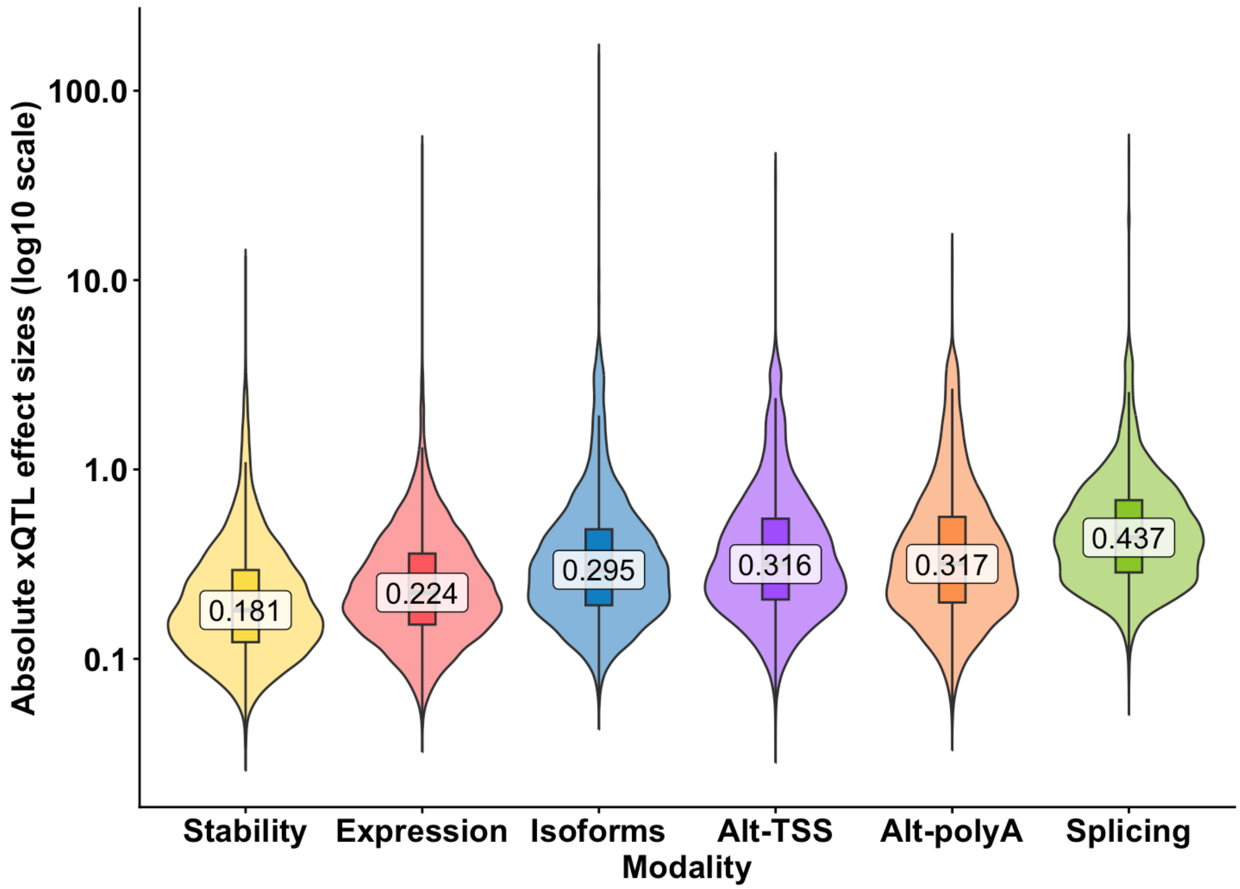


**Figure S2. Distributions of the absolute effect sizes of conditionally independent xQTLs per gene in six transcriptional modalities.**

Violin plots with overlaid boxplots on a log10 scale comparing the absolute effect size distributions across six transcriptional modalities. We used all conditionally independent xQTLs identified for each gene using separate-modality mapping.

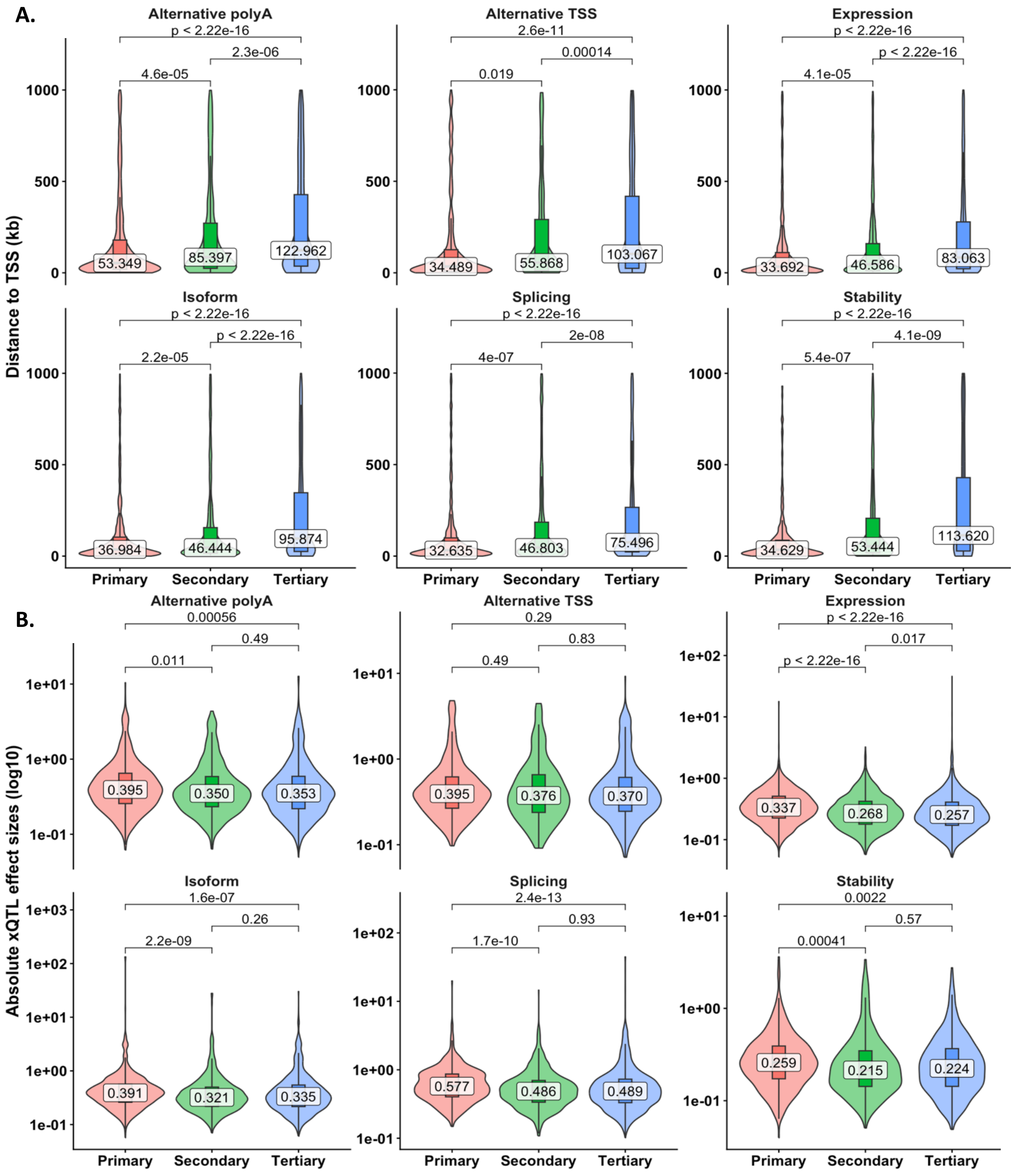


**Figure S3. Characteristics of independent xQTL signals within each modality for primary, secondary, and tertiary+ xQTLs in xGenes that contained 3+ independent xQTLs.**

(A) The distribution of distances to the corresponding TSS; (B) the distribution of absolute effect sizes. Numbers inside the boxplots indicate median absolute effect sizes. P-values from pairwise Wilcoxon rank-sum tests are shown for each plot.

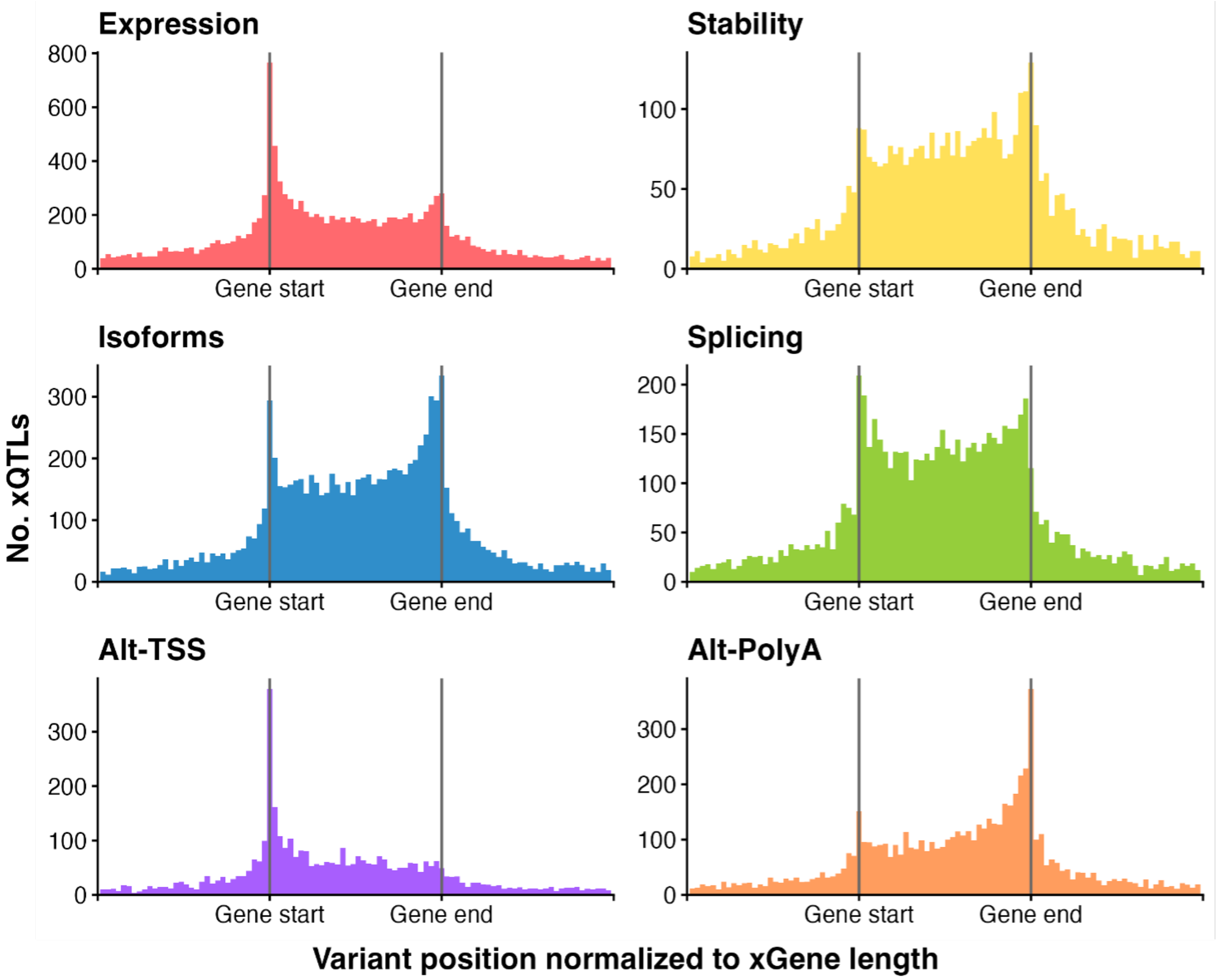


**Figure S4. Positional distributions of xQTLs mapped by separate-modality analyses.**

QTL base pair positions were scaled by gene length to the same TSS (gene start) and TES positions (gene end). The plot displays only xQTLs located within the window of one gene's length around the gene TSS or TES.

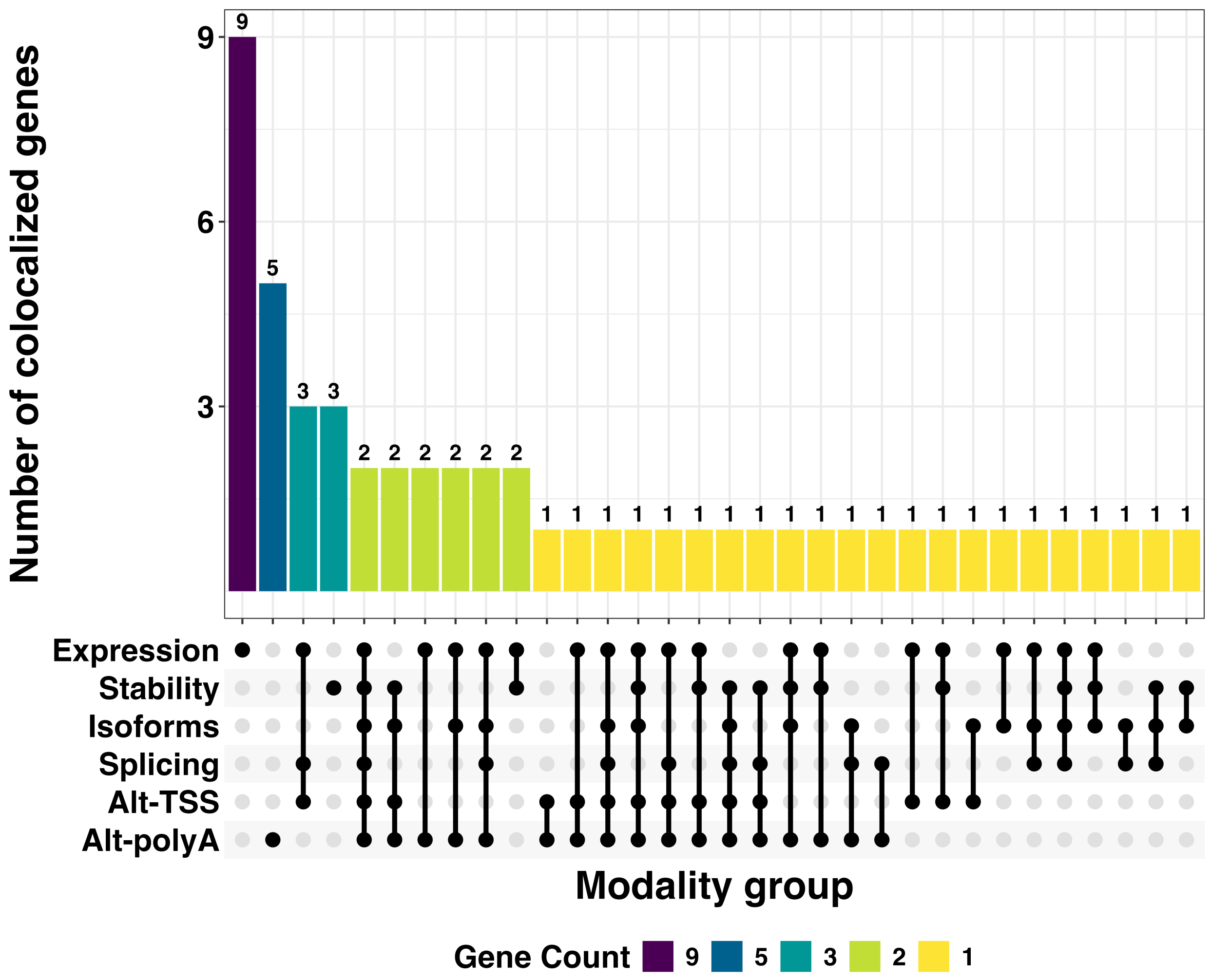


**Figure S5. Upset plot of AD GWAS risk genes colocalized in six modalities by separate-modality analysis.**

Bar color corresponds to the number of colocalized AD GWAS risk xGenes. The vertical bars represent the number of colocalized xGenes within each specific intersections of modalities, as denoted by the connected dots in the bottom.

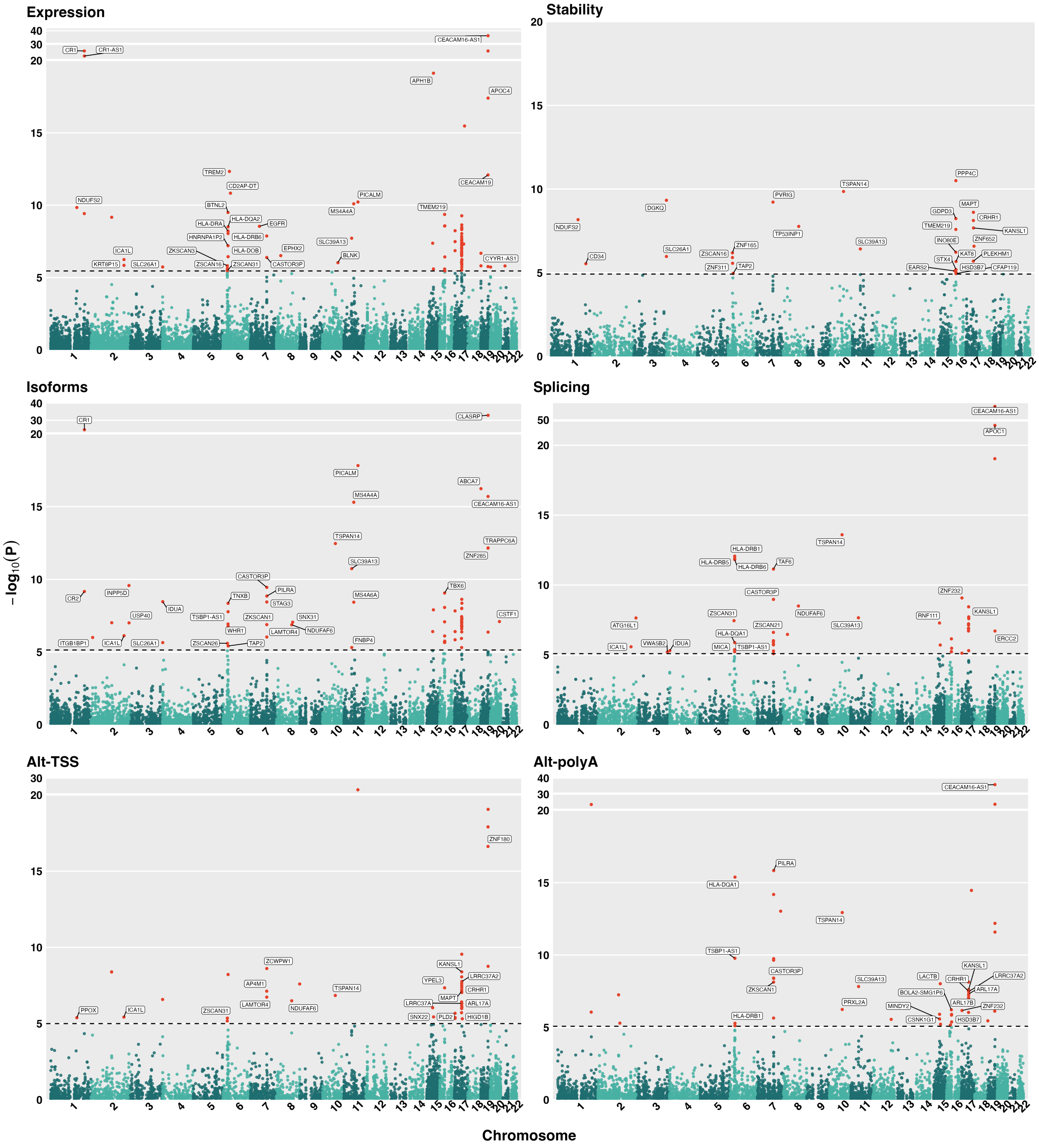


**Figure S6. Manhattan plots of xTWAS results by FUSION/BestModel per modality.**

The -log10(xTWAS p-values) are plotted on the y-axis, versus gene TSS positions on the x-axis. The dashed line represents the genome-wide significance threshold after Bonferroni correction. Significant xTWAS risk genes with available HGNC symbols are labeled. Some labels may be omitted due to the max.overlaps setting in the R ggrepel package.

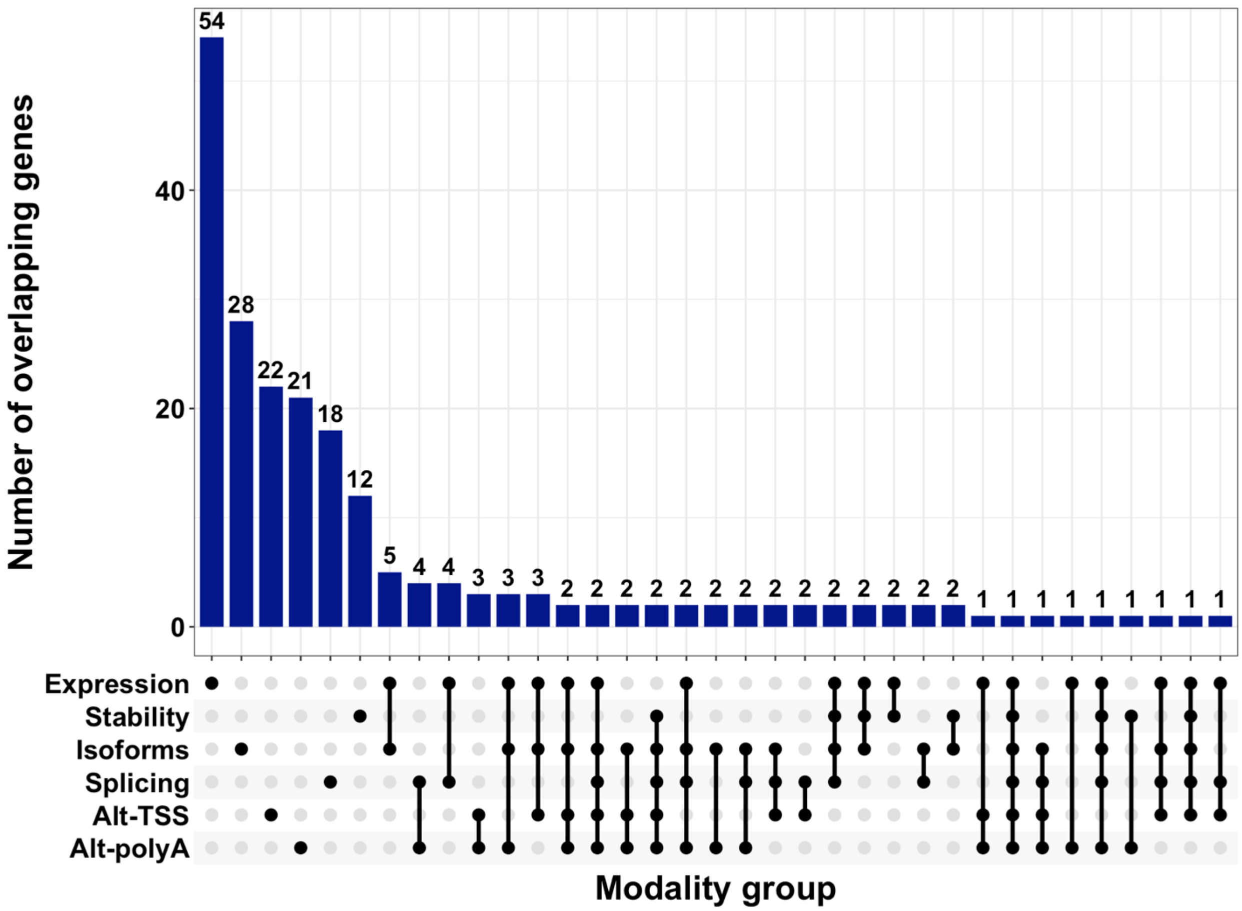


**Figure S7. Upset plot of all xTWAS risk genes of AD in six transcriptional modalities.** The vertical bars represent the number of TWAS risk genes within each specific intersections of modalities, as denoted by the connected dots in the bottom.

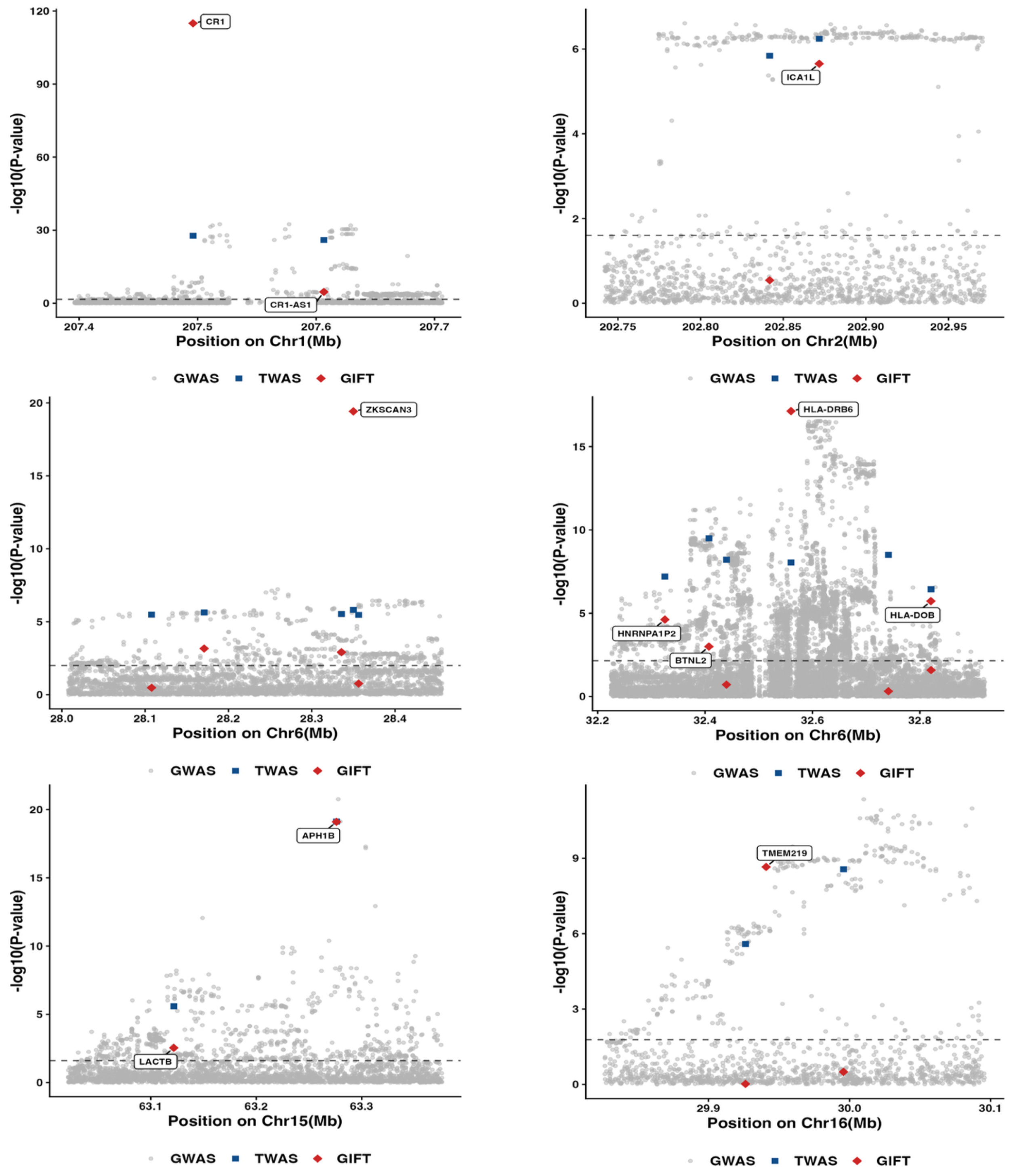


**Figure S8. Locus-zoom plots of fine-mapped regions with at least one significant independent xTWAS risk gene in the expression modality by GIFT (in Chromosomes 1, 2, 6, 15, and 16).**

SNPs by GWAS, genes identified by xTWAS, and GIFT were plotted in grey dots, blue squares, and red diamonds, respectively. The Bonferroni corrected threshold of each test region was used to identify significant fine-mapped genes by GIFT (dashed horizontal line). Fine-mapped independent significant genes by GIFT are labelled.

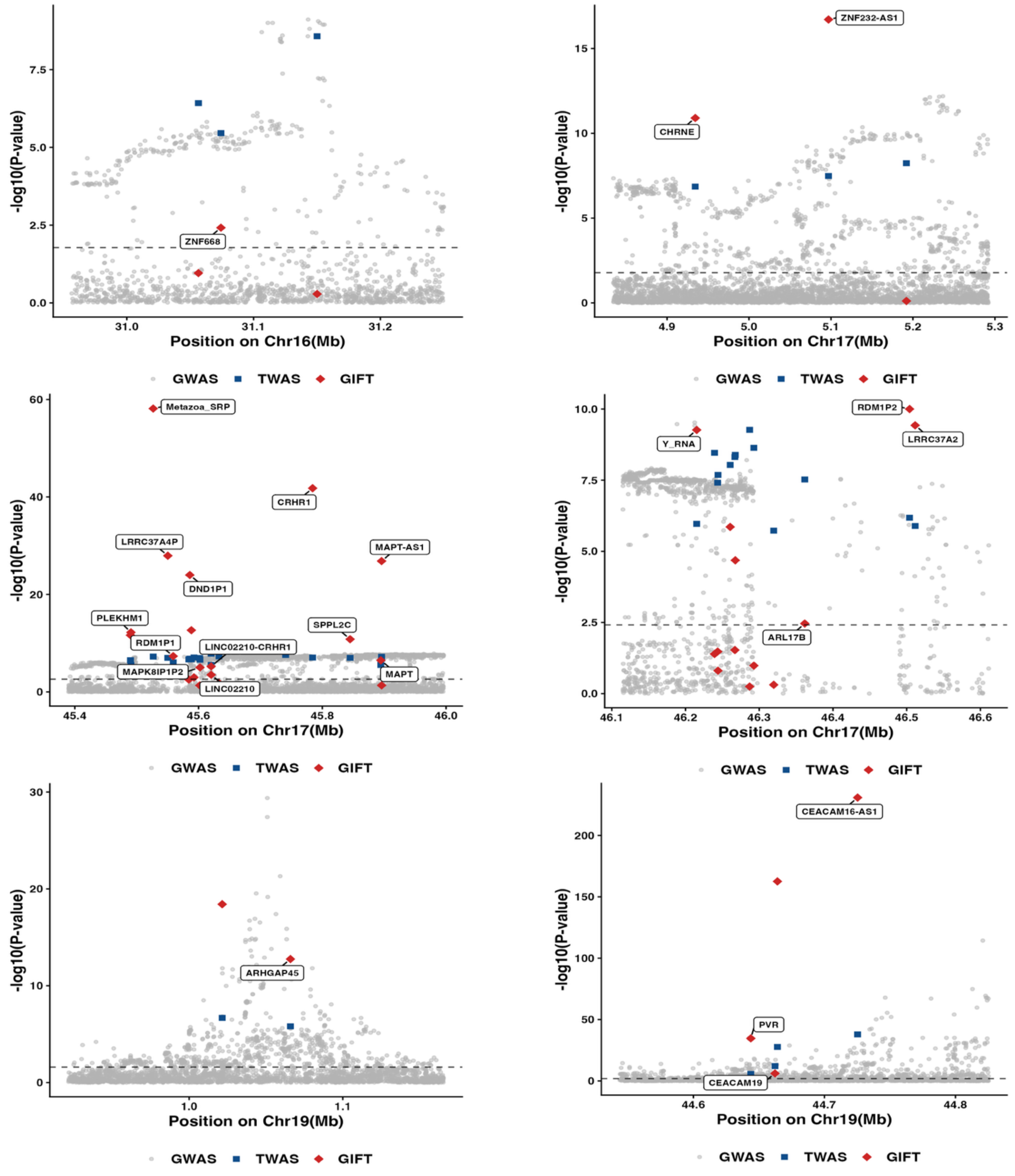


**Figure S9. Locus-zoom plots of fine-mapped regions with at least one significant independent TWAS risk gene in the expression modality by GIFT (Chromosomes 16, 17, and 19).**

SNPs by GWAS, genes identified by xTWAS, and GIFT were plotted in grey dots, blue squares, and red diamonds, respectively. The Bonferroni corrected threshold of each test region was used to identify significant fine-mapped genes by GIFT (dashed horizontal line). Fine-mapped independent significant genes by GIFT are labelled.

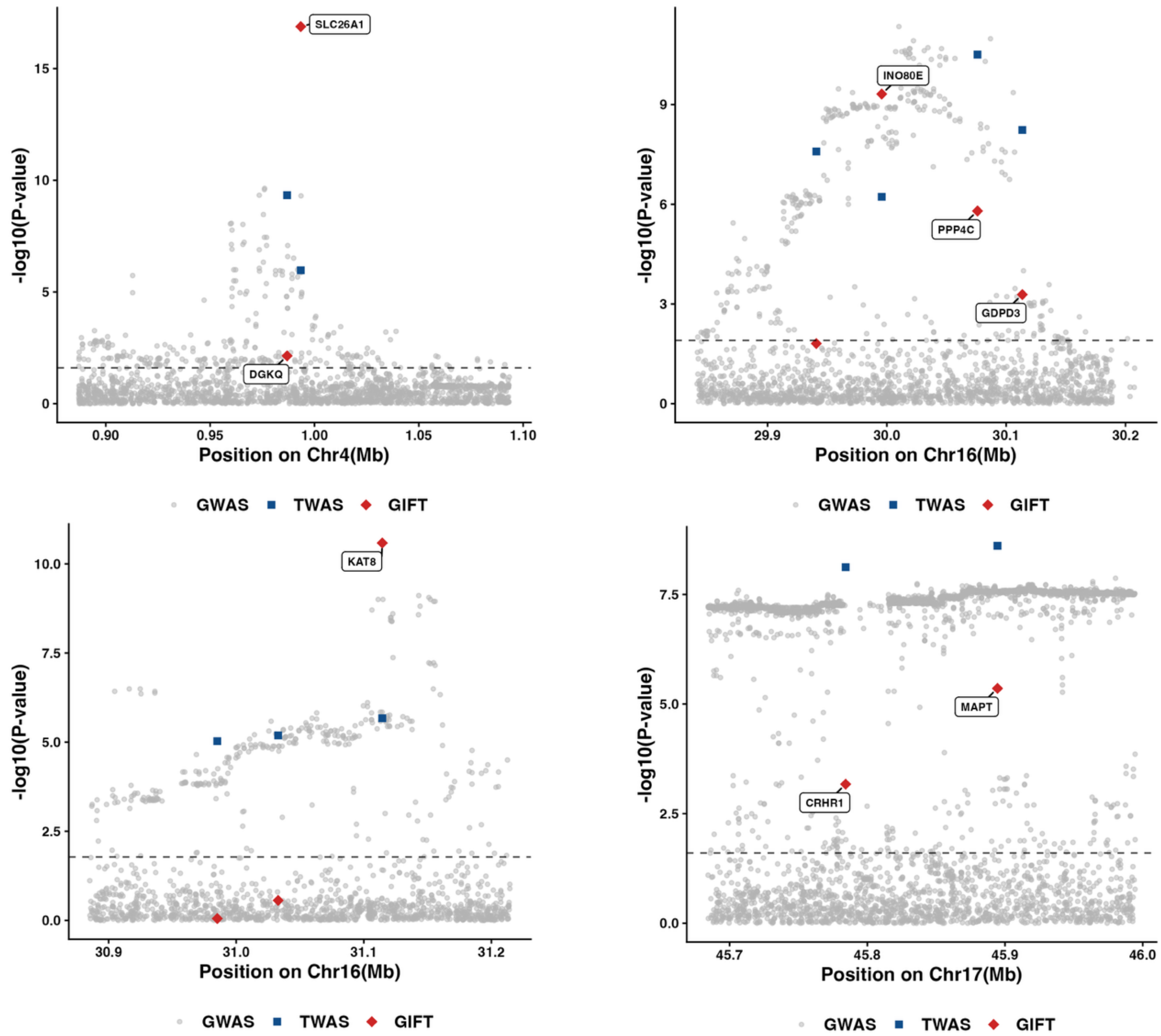


**Figure S10. Locus-zoom plots of fine-mapped regions with at least one significant risk gene by GIFT in Stability.**

SNPs by GWAS, genes identified by xTWAS, and GIFT were plotted in grey dots, blue squares, and red diamonds, respectively. The Bonferroni corrected threshold of each test region was used to identify significant fine-mapped genes by GIFT (dashed horizontal line). Fine-mapped independent significant genes by GIFT are labelled.

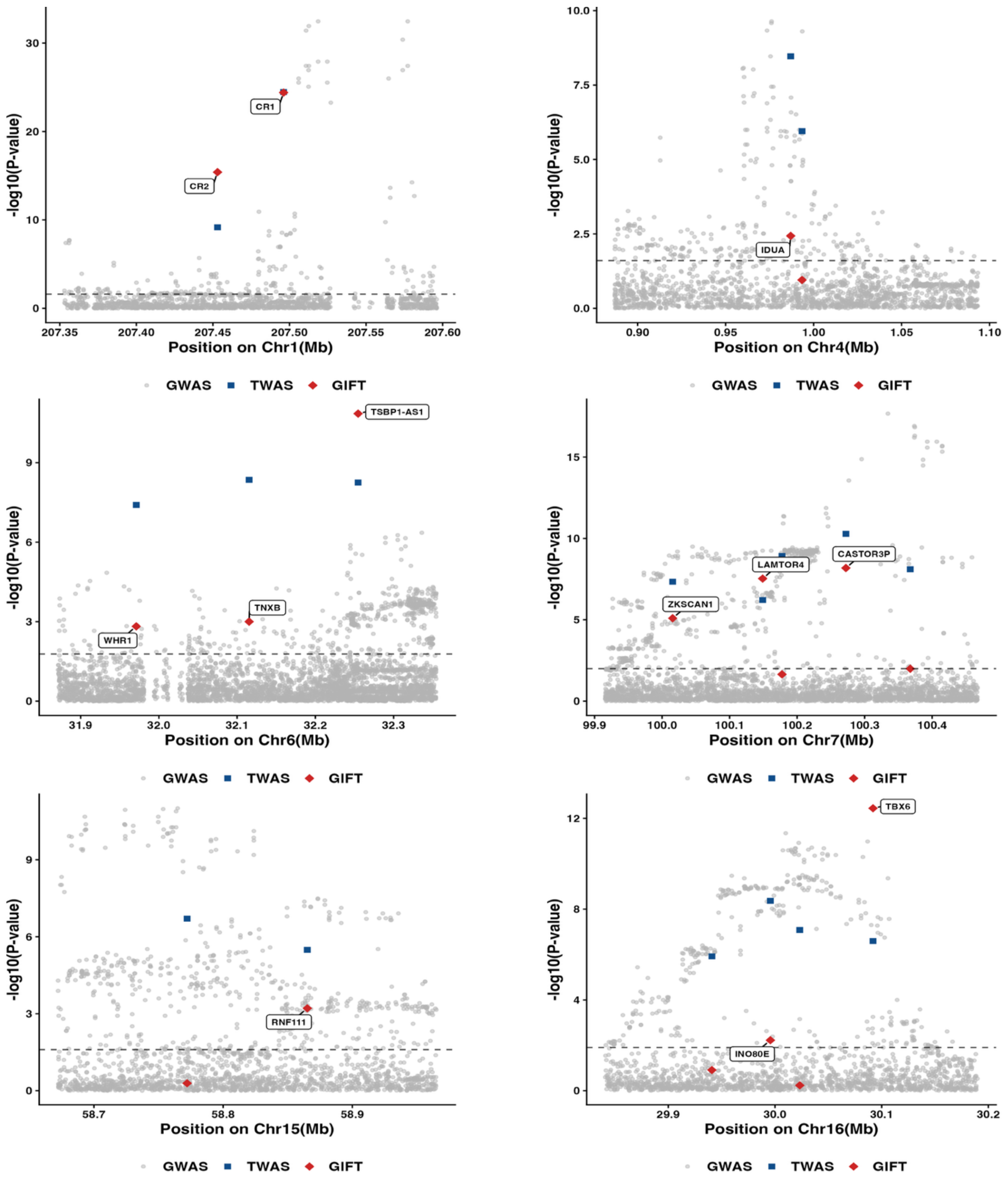


**Figure S11. Locus-zoom plots of fine-mapped regions with at least one significant risk gene by GIFT in Isoforms (Chromosomes 1, 4, 6, 7, 15, and 16).**

SNPs by GWAS, genes identified by xTWAS, and GIFT were plotted in grey dots, blue squares, and red diamonds, respectively. The Bonferroni corrected threshold of each test region was used to identify significant fine-mapped genes by GIFT (dashed horizontal line). Fine-mapped independent significant genes by GIFT are labelled.

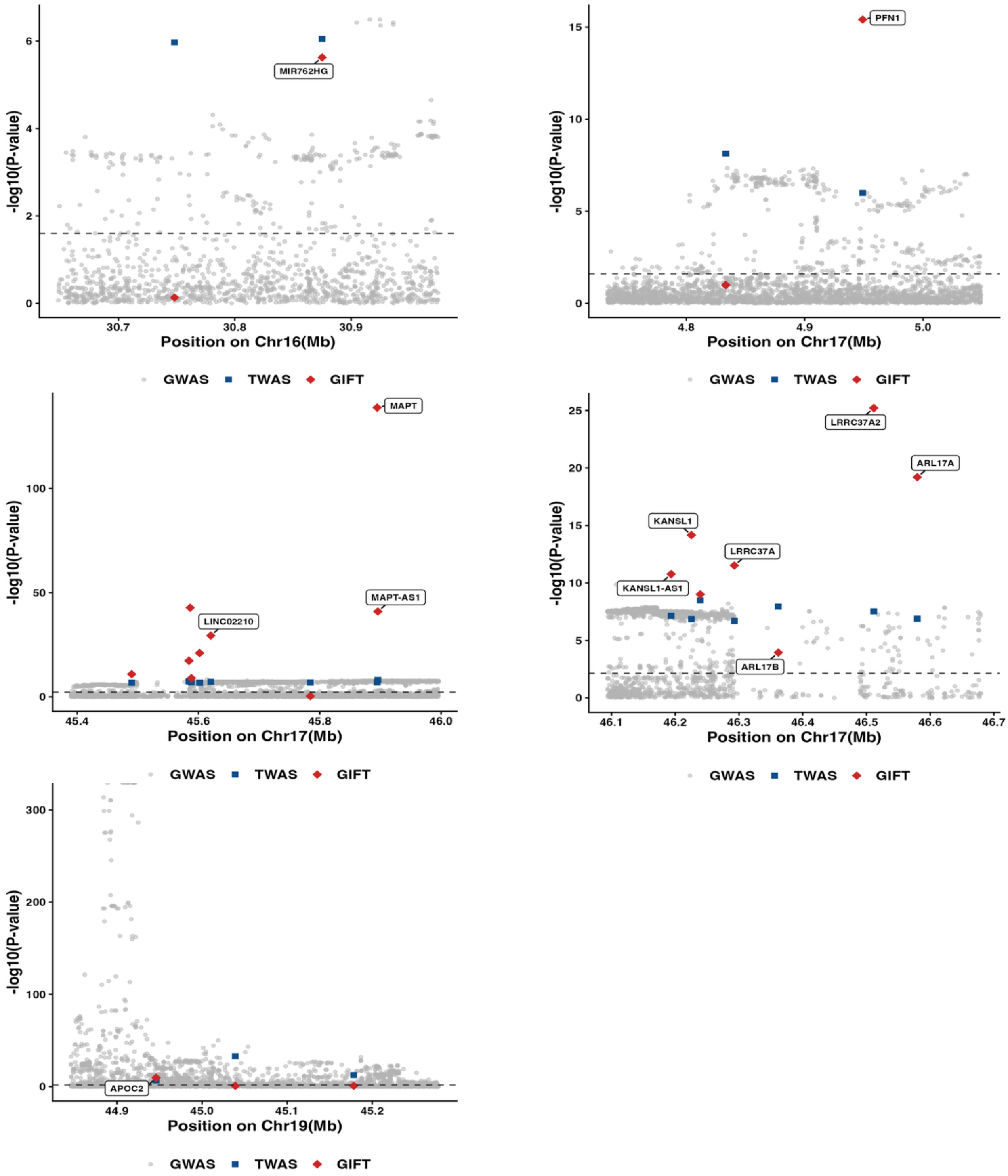


**Figure S12. Locus-zoom plots of fine-mapped regions with at least one significant risk gene by GIFT in Isoforms (Chromosome 16, 17, 19).**

SNPs by GWAS, genes identified by xTWAS, and GIFT were plotted in grey dots, blue squares, and red diamonds, respectively. The Bonferroni corrected threshold of each test region was used to identify significant fine-mapped genes by GIFT (dashed horizontal line). Fine-mapped independent significant genes by GIFT are labelled.

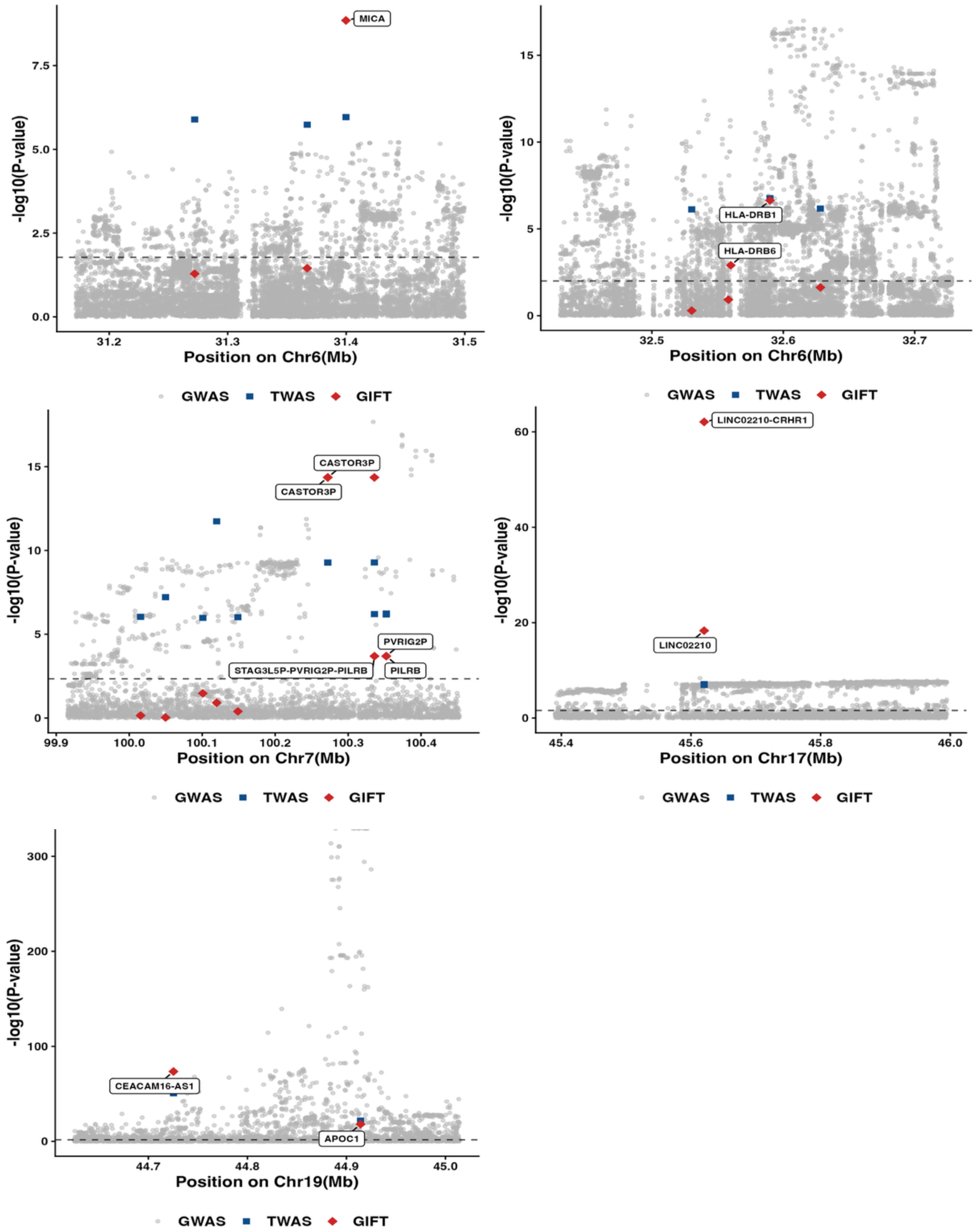


**Figure S13. Locus-zoom plots of fine-mapped regions with at least one significant risk gene by GIFT in Splicing.**

SNPs by GWAS, genes identified by xTWAS, and GIFT were plotted in gray dots, blue squares, and red diamonds, respectively. The Bonferroni corrected threshold of each test region was used to identify significant fine-mapped genes by GIFT (dashed horizontal line). Fine-mapped independent significant genes by GIFT are labelled.

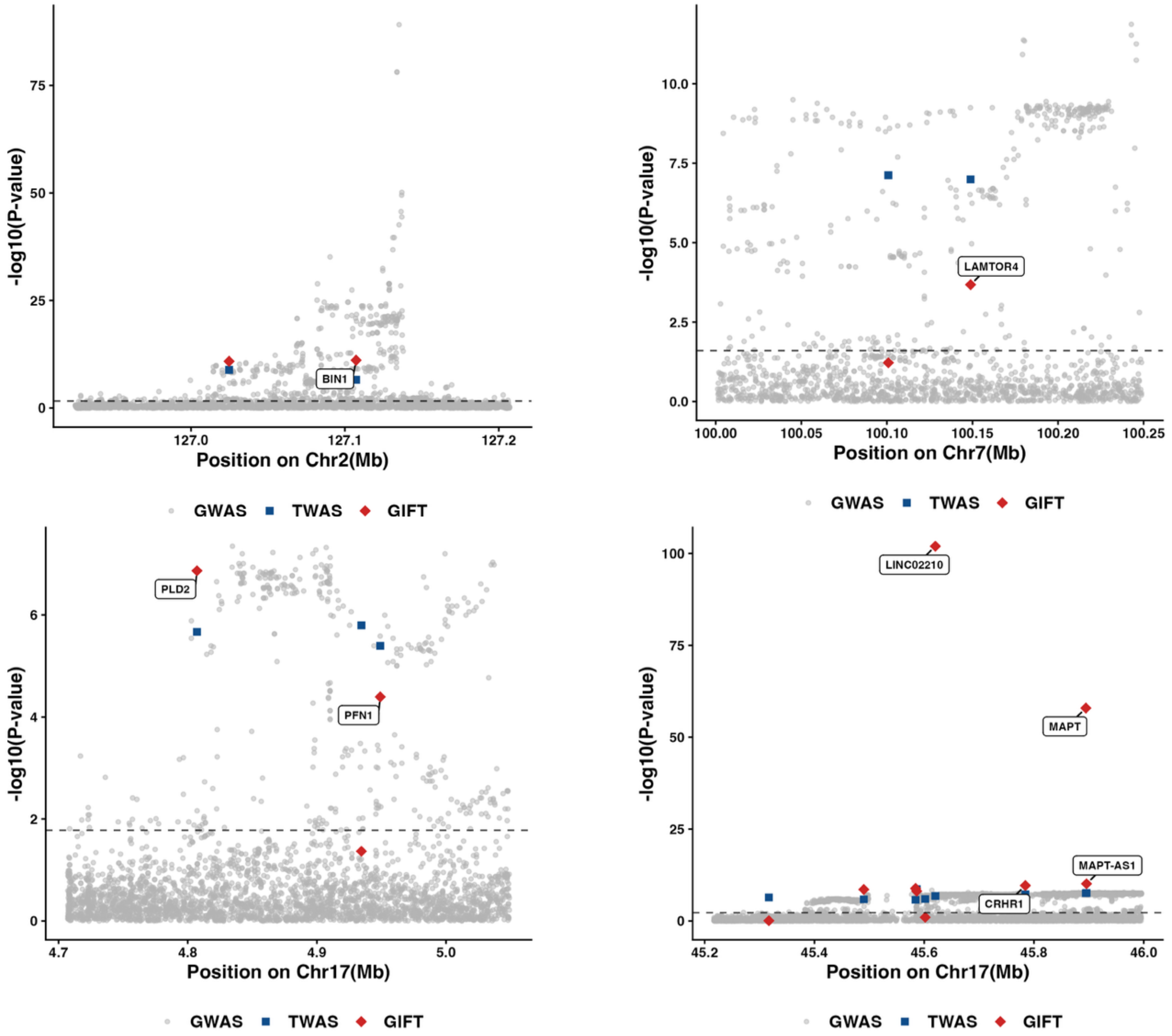


**Figure S14. Locus-zoom plots of fine-mapped regions with at least one significant risk gene by GIFT in Alternative TSS (Chromosome 2, 7, and 17).**

SNPs by GWAS, genes identified by xTWAS, and GIFT were plotted in grey dots, blue squares, and red diamonds, respectively. The Bonferroni corrected threshold of each test region was used to identify significant fine-mapped genes by GIFT (dashed horizontal line). Fine-mapped independent significant genes by GIFT are labelled.

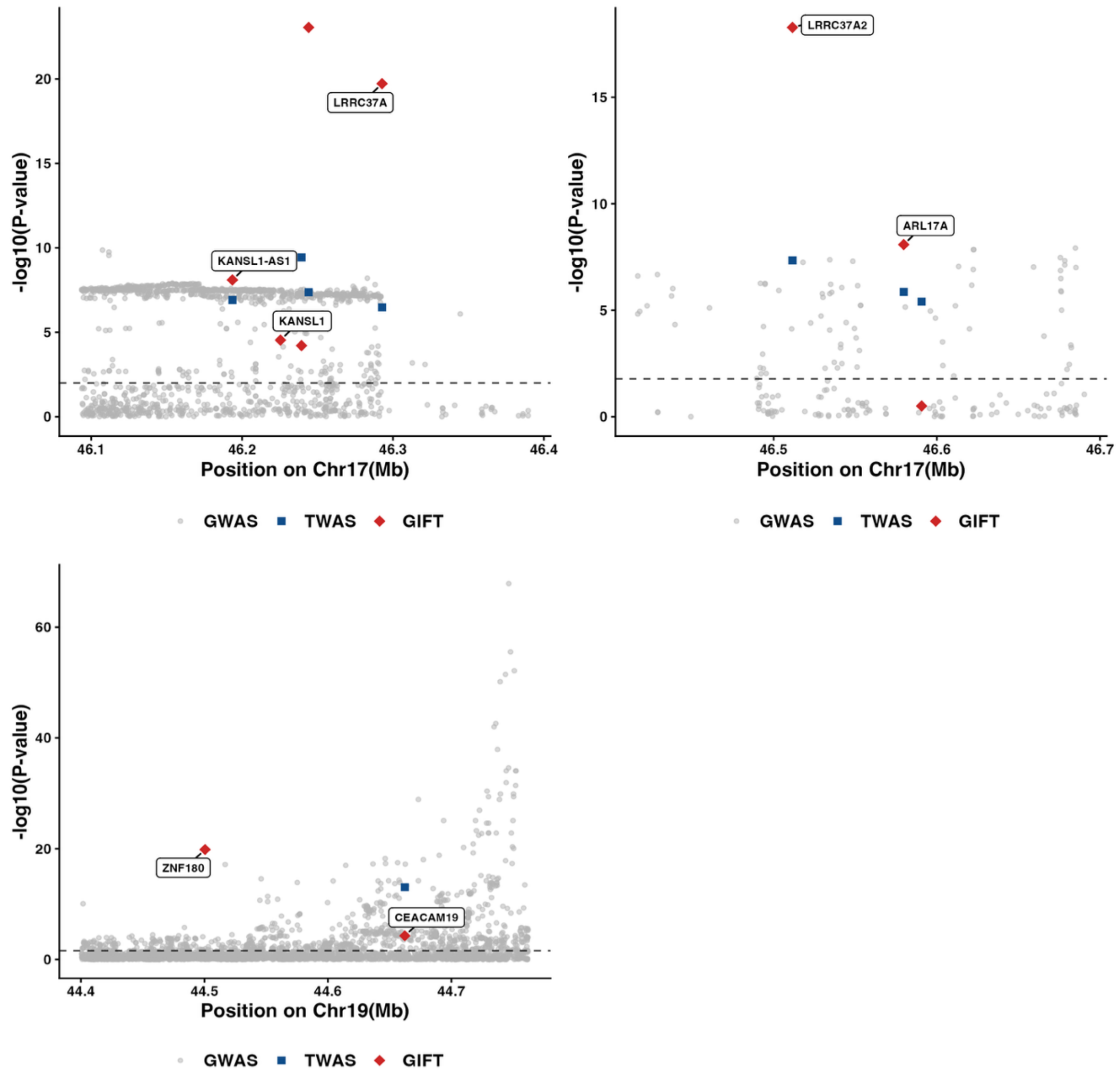


**Figure S15. Locus-zoom plots of fine-mapped regions with at least one significant risk gene by GIFT in Alternative TSS (Chromosome 17 and 19).**

SNPs by GWAS, genes identified by xTWAS, and GIFT were plotted in gray dots, blue squares, and red diamonds, respectively. The Bonferroni corrected threshold of each test region was used to identify significant fine-mapped genes by GIFT (dashed horizontal line). Fine-mapped independent significant genes by GIFT are labelled.

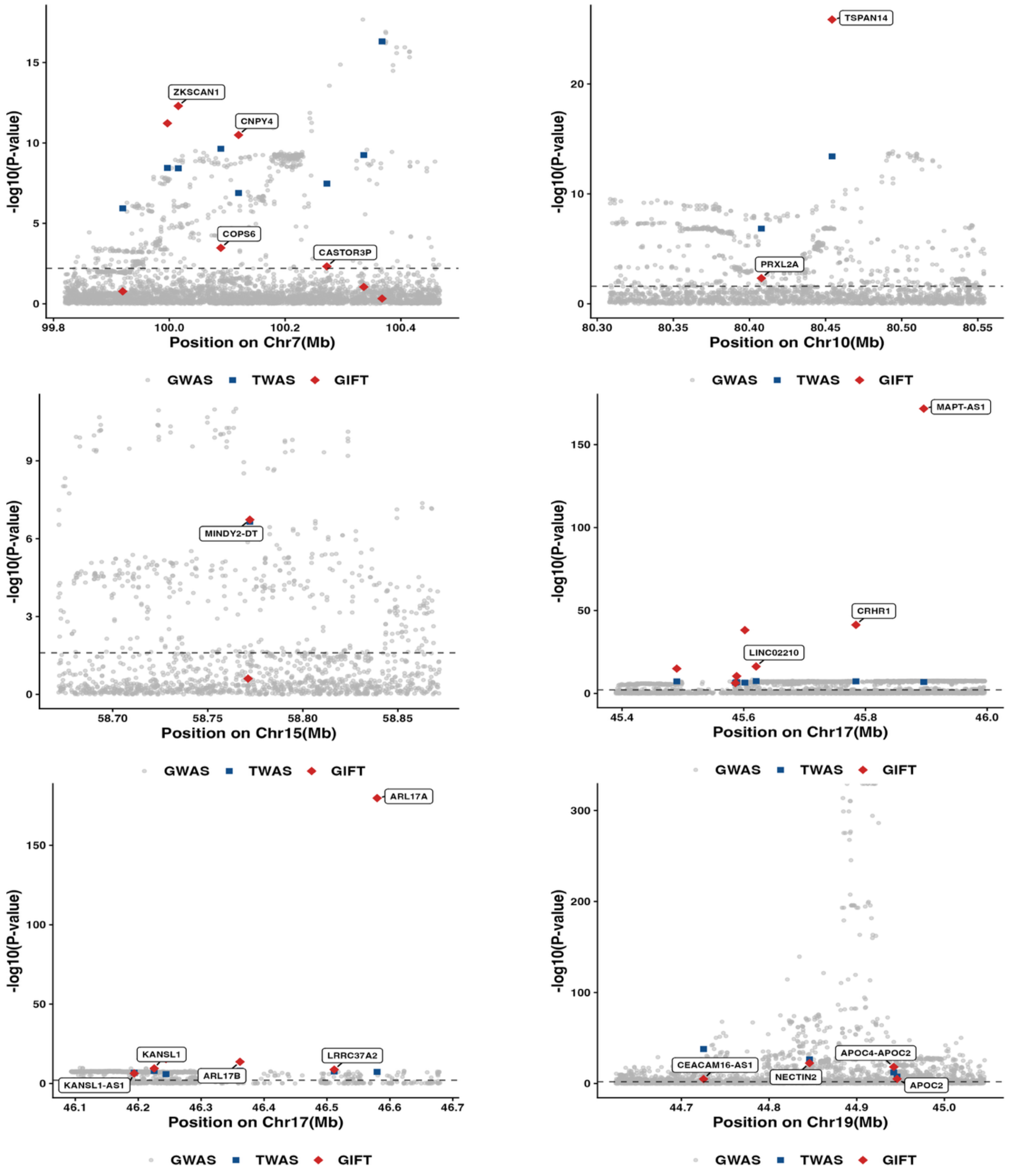


**Figure S16. Locus-zoom plots of fine-mapped regions with at least one significant risk gene by GIFT in Alternative polyA.**

SNPs by GWAS, genes identified by TWAS, and GIFT were plotted in grey dots, blue squares, and red diamonds, respectively. The Bonferroni corrected threshold of each test region was used to identify significant fine-mapped genes by GIFT (dashed horizontal line). Fine-mapped independent significant genes by GIFT are labelled.

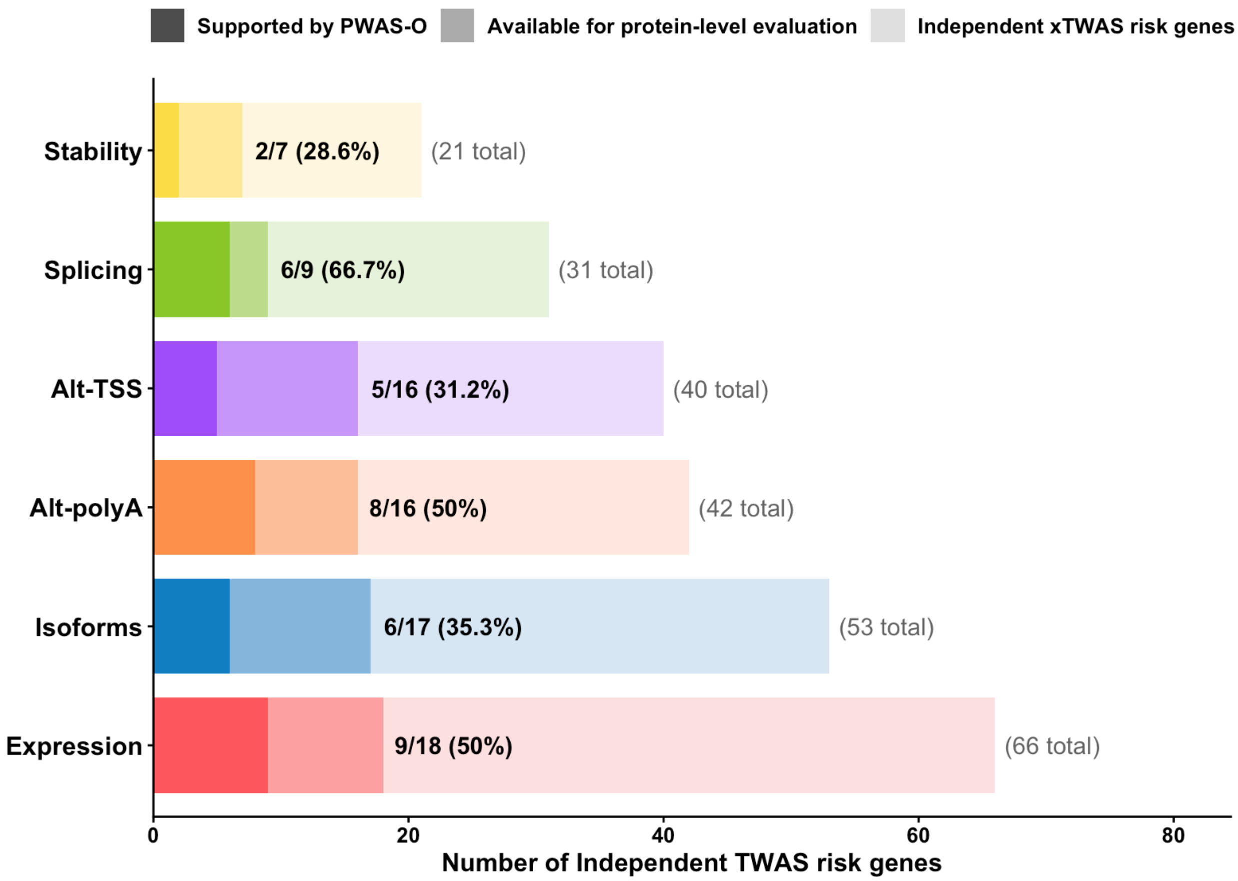


**Figure S17. Proportions of independent xTWAS risk genes of AD validated in proteomics in six transcriptomic modalities.**

For each modality, the total number of independent xTWAS risk genes is shown by the lightest shade, with the total number displayed to the right of each bar (e.g., 66 total). The medium shade indicates genes available for protein-level evaluation based on genomic region overlap with genes tested by PWAS-O, and the darkest shade indicates xTWAS risk genes further supported by significant PWAS-O risk genes (q-value < 0.05). Labels within the bars indicate the number and percentage of genes supported by PWAS-O among those available for protein-level evaluation (supported/available, %).

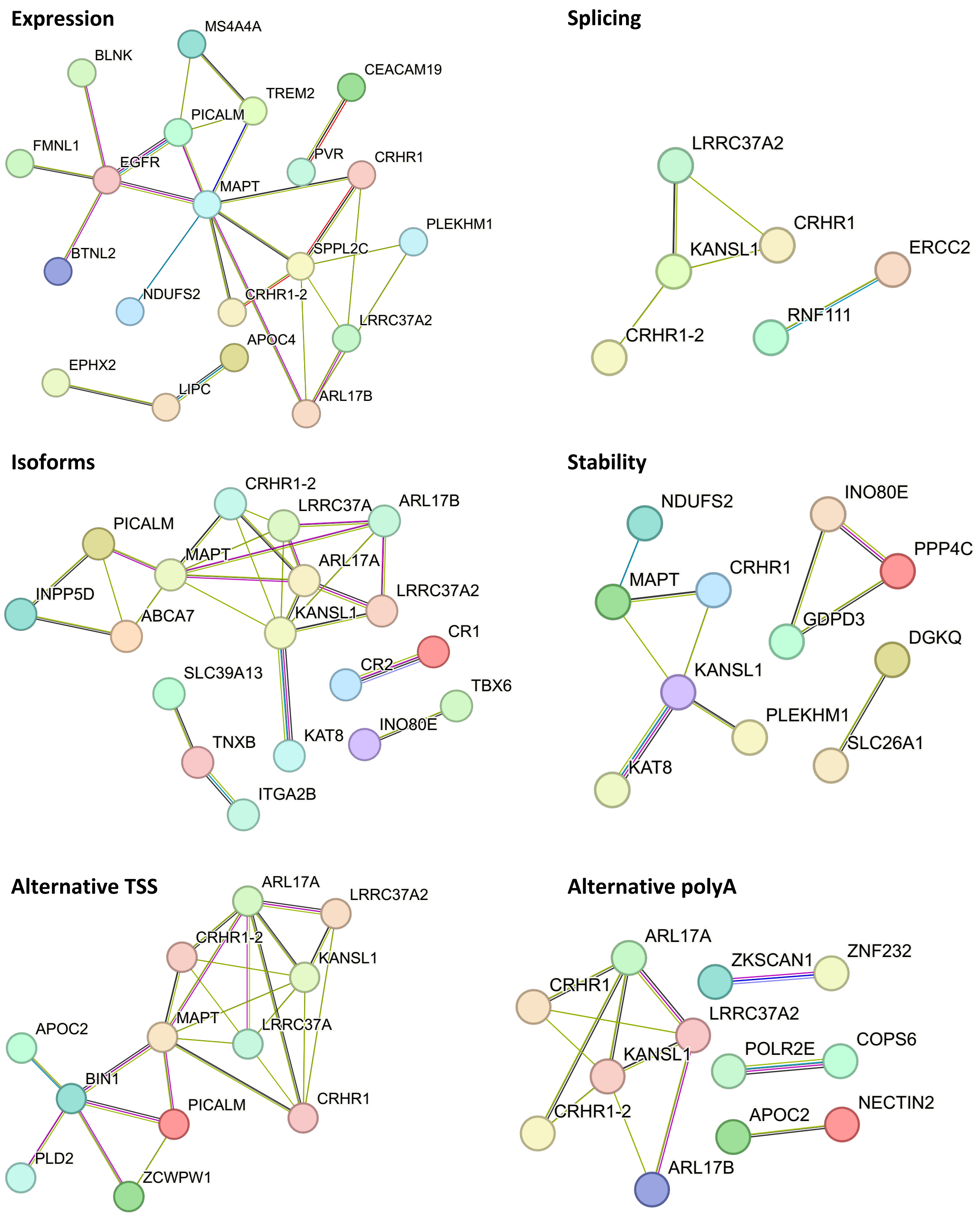


**Figure S18. PPI networks of independent xTWAS significant genes of AD within each modality.** Edges represent PPI links, with different colors representing different sources of connection evidence. Disconnected nodes are excluded from the plots.

| Result | Gene expression | RNA stability | Isoform ratios | Splice junction usage | Alternative TSS usage | Alternative polyA usage | Unique across modalities |
|---|---|---|---|---|---|---|---|
| **Independent xQTLs (Separate)** | 26,131 | 9,714 | 14,850 | 11,492 | 6,723 | 11,453 | **80,363** |
| **xGenes (Separate)** | 18,143 | 6,790 | 9,111 | 6,902 | 5,032 | 7,757 | **25,736** |
| **Independent xQTLs (Cross)** | 19,024 | 3,230 | 5,632 | 7,150 | 3,787 | 5,734 | **44,557** |
| **xGenes (Cross)** | 14,149 | 2,790 | 4,227 | 4,499 | 3,029 | 4,447 | **23,811** |
| **Colocalized GWAS loci (Separate)** | 35 | 20 | 21 | 17 | 18 | 27 | **54** |
| **Colocalized GWAS loci (Cross)** | 40 | 14 | 17 | 12 | 9 | 15 | **55** |
| **xTWAS risk genes** | 88 | 26 | 68 | 48 | 46 | 51 | **214** |
| **Independent xTWAS risk genes** | 66 | 21 | 53 | 29 | 40 | 42 | **161** |
| **PWAS-O support** | 9 | 2 | 6 | 6 | 5 | 8 | **26** |
| **PPI network membership (% out of independent xTWAS risk genes)** | 20 (30.3%) | 11 (52.4%) | 18 (34.0%) | 6 (20.7%) | 12 (30.0%) | 12 (28.6%) | **75 (46.6%)** |
| **Enriched Pathway** | T cell activation | Wnt signaling, pathway | Integrin signaling, Glycolysis | Blood coagulation | Integrin signaling, Axon guidance, Synaptic vesicle trafficking | Toll-Like Receptor, Intracellular signaling, Complement pathway | — |

**Figure S19. Summary of key findings across six transcriptomic modalities.**

The table summarizes the number so conditionally independent xQTLs, xGenes identified through separate- and cross-modality mapping, colocalized AD GWAS loci, significant xTWAS genes, and independent xTWAS risk genes for each modality. The rightmost column shows unique counts across all six modalities rather than sums of modality-specific counts. PWAS-O support indicates the number of independent xTWAS risk genes supported by proteomic data. PPI network membership indicates the number of independent xTWAS risk genes connected in STRING PPI interaction networks. Enriched pathways indicate the number of significantly enriched pathways for each modality.

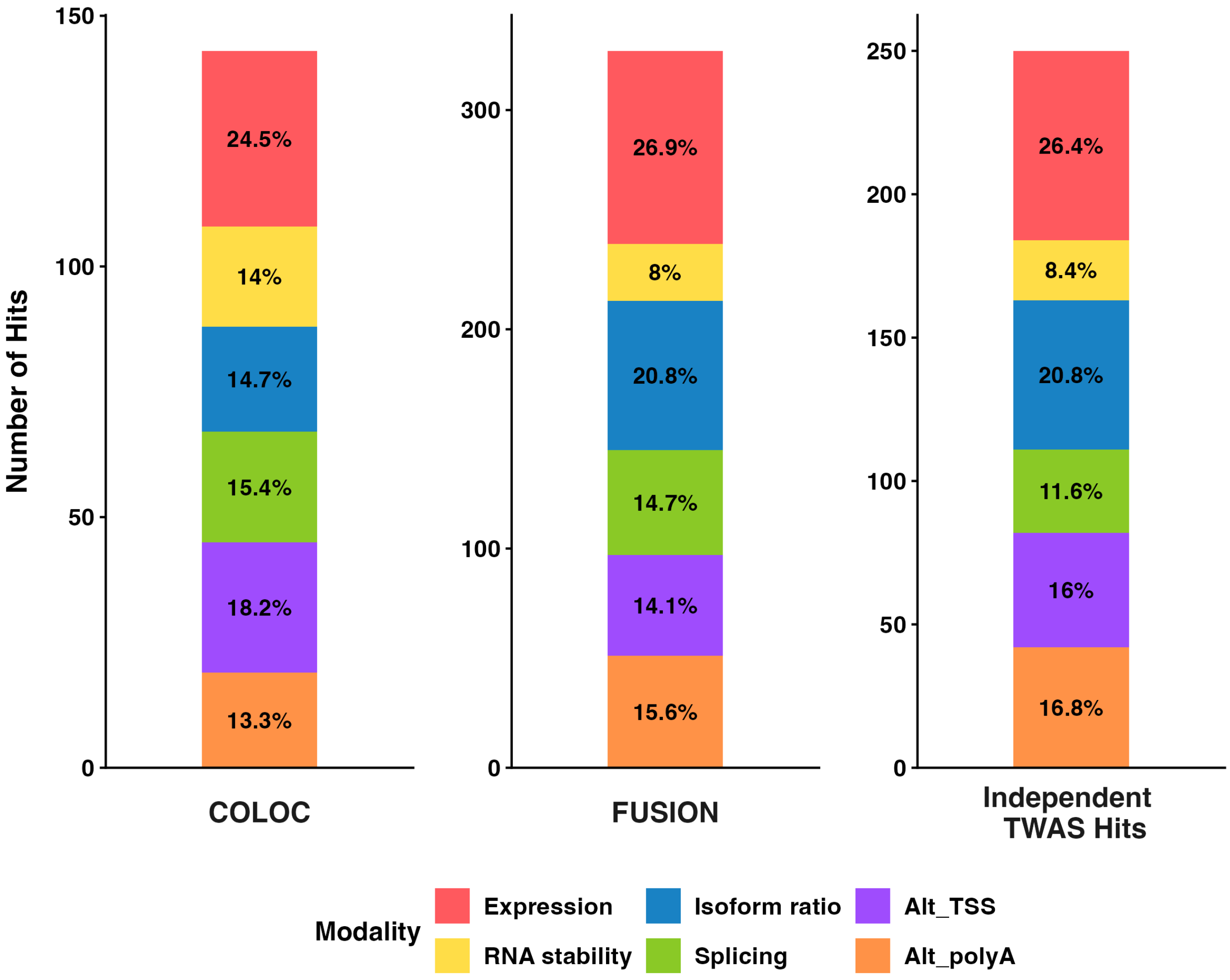


**Figure S20. Proportion of modalities contributing significant hits to the findings by colocalization, FUSION, and independent xTWAS analyses.**

The total number of hits plotted on the y-axis contain duplicate genes that are identified in multiple modalities.

**A. Isoform ratio**

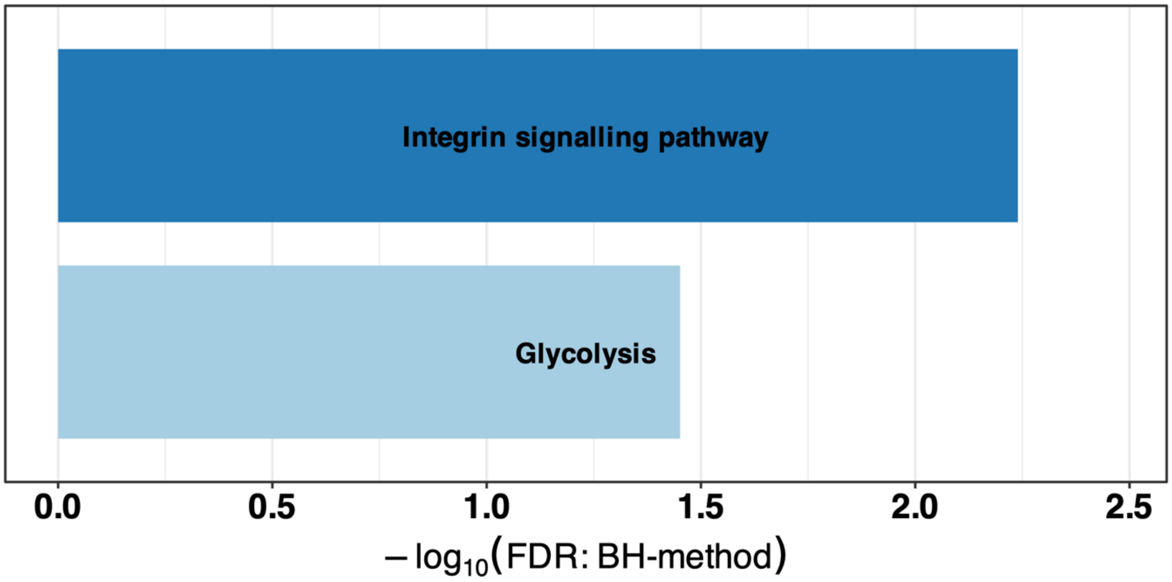


**B. Alternative TSS usage**

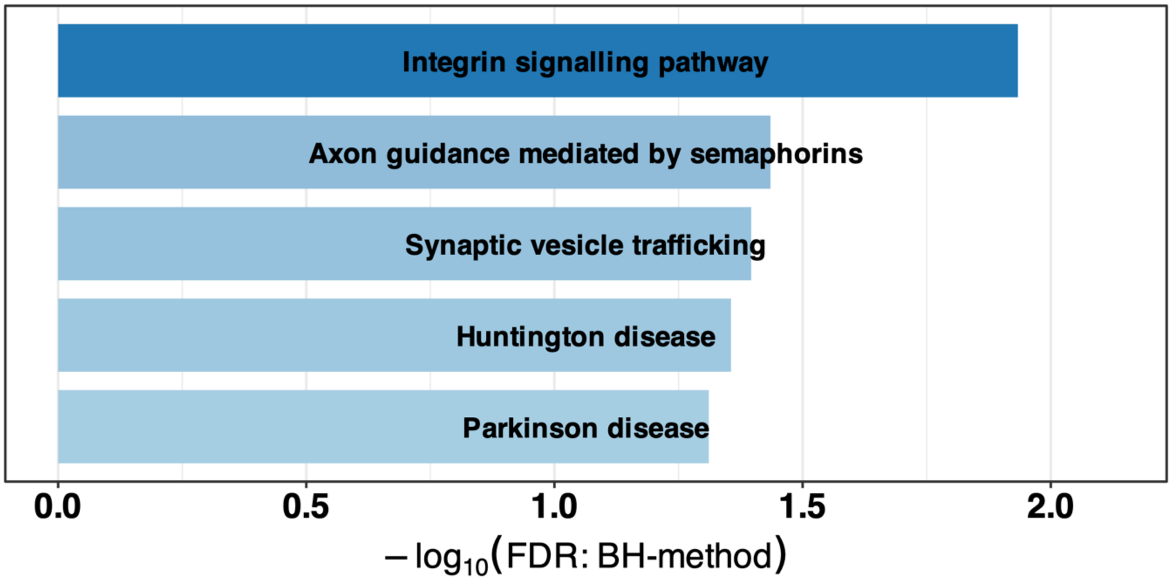


**Figure S21. Examples of significant gene pathways enriched with xTWAS risk genes identified in non-expression modalities (isoform ratio and alternative TSS usage).**

Pathway significantly enriched (FDR $q < 0.05$) among AD xTWAS risk genes identified in (A) isoform ratio and (B) alternative TSS usage. The x-axis represents the log10-transformed Benjamini-Hochberg adjusted p-values.

### Supplementary Tables

**For supplemental data 1-9, see attached supplemental spreadsheet.**

| Transcriptional modality | No. RNA traits | No. genes with RNA traits | No. RNA traits per gene (mean ± SD) |
|---|---|---|---|
| Gene expression | 50,735 | 50,735 | 1 ± 0 |
| RNA Stability | 12,436 | 12,436 | 1 ± 0 |
| Isoforms Ratio | 56,383 | 20,384 | 2.77 ± 1.66 |
| Splice Junction Usage | 103,103 | 17,499 | 5.91 ± 4.65 |
| Alternative TSS Usage | 89,085 | 22,960 | 3.89 ± 2.25 |
| Alternative PolyA Usage | 84,602 | 23,193 | 3.65 ± 2.16 |

**Table S1. Characteristics of quantified RNA traits of six transcriptional modalities.**

| Modality | Conditionally Independent xQTLs | No. xGenes |
|---|---|---|
| Gene Expression | 26,131 | 18,143 |
| RNA Stability | 9,714 | 6,790 |
| Isoform Ratio | 14,850 | 9,111 |
| Splice Junction Usage | 11,492 | 6,902 |
| Alternative TSS Usage | 6,723 | 5,032 |
| Alternative PolyA Usage | 11,453 | 7,757 |

**Table S2. Summary Statistics of conditionally independent xQTLs and the number of xGenes by separate-modality xQTL analyses.**

| Modality | Colocalized GWAS loci with separate-modality xQTL | Colocalized GWAS loci with cross-modality xQTL |
|---|---|---|
| Gene Expression | 35 (46.7%) | 40 (53.3%) |
| RNA Stability | 20 (26.7%) | 14 (18.7%) |
| Isoforms Ratio | 21 (28%) | 17 (22.7%) |
| Splice Junction Usage | 17 (22.7%) | 12 (16%) |
| Alternative TSS Usage | 18 (24%) | 9 (12%) |
| Alternative PolyA Usage | 27 (36%) | 15 (20%) |

**Table S3. The number of AD GWAS risk genes that colocalized with xQTL signals in six transcriptional modalities by separate- and cross-modality analyses.**

Values represent the number of colocalized GWAS loci and the percentage out of the 75 analyzed AD GWAS risk loci, using a posterior probability ($PP_{H4}$) threshold of 0.75. The second and third columns correspond to separate-modality and cross-modality xQTL analyses, respectively.

| Modality | No. Tested RNA Traits | Median CV $R^2$ | No. Tested Genes | Significant xTWAS Genes |
|---|---|---|---|---|
| Gene Expression | 14,218 | 0.029 | 14,218 | 88 |
| RNA Stability | 4,161 | 0.028 | 4,161 | 26 |
| Isoform Ratio | 13,216 | 0.026 | 6,945 | 68 |
| Splice Junction Usage | 17,713 | 0.022 | 6,128 | 48 |
| Alternative TSS | 11,632 | 0.014 | 4,937 | 46 |
| Alternative PolyA | 14,698 | 0.021 | 5,925 | 51 |

**Table S4**. **Results of xTWAS analyses of AD by FUSION.**

The number of RNA traits analyzed by FUSION are shown in the 2nd column. RNA traits that met the significant cis-SNP heritability criterion (LRT p-value < 0.01) were considered as tested RNA traits, with the model having the highest cross-validation (CV) $R^2$ selected among Top1, BLUP, LASSO, and Elastic Net. The median cross validation (CV) $R^2$ given by the best-selected prediction model of tested RNA traits are shown in the 3rd column. The number of unique genes with the tested RNA traits are shown in the 4th column. The number of significant genes (with Bonferroni adjusted p-values <0.05) are shown in the 5th column, where xTWAS p-values of RNA traits are aggregated by ACAT per gene for modalities with multiple RNA traits per gene (isoform ratio, splice junction usage, alternative TSS, and alternative polyA).

| Modality | Independent significant genes by GIFT | Independent significant genes not overlapped with any others | Total independent TWAS risk genes |
|---|---|---|---|
| Gene Expression | 44 | 22 | 66 |
| RNA Stability | 8 | 13 | 21 |
| Isoform Ratio | 30 | 23 | 53 |
| Splice Junction Usage | 12 | 17 | 29 |
| Alternative TSS | 22 | 18 | 40 |
| Alternative PolyA | 25 | 17 | 42 |

**Table S5**. **Number of independent xTWAS risk genes of AD.**

### Supplemental Methods

#### Quantification of RNA traits of six transcriptomic modalities

The following six transcriptomic modalities were quantified using the corresponding existing state-of-art RNA data analysis tools that were employed within the Snakemake[1] architecture of the Pantry[2] framework.

**Total gene expression and Isoform ratio**: Transcript-level abundances were quantified in transcript per million (TPM) units using kallisto[3]. Total gene expression was computed by summing isoform-level read abundances across all transcripts mapped to a specific gene. For the isoform ratio modality, isoform-level read abundances were normalized per gene to determine the relative proportion of alternatively spliced transcript (i.e. isoforms) from a single gene.

**RNA stability**: Relative steady-state stability of RNA molecules was measured by calculating the ratio of mature to pre-mRNA abundance. Subread-featureCounts[4] were used to count reads from constitutive exons and all introns, then the exon-to-intron read ratio per gene was calculated to estimate relative RNA stability. To quantify RNA stability, gene annotations were filtered by Pantry to include only those with the Ensembl pipeline as the annotation source, thereby limiting rare or speculative isoforms that would prevent any constitutive exons from being counted across many genes.

**Splice junction usage (Intron excision ratio)**: Variation in mRNA splicing events was captured by measuring the relative rates at which specific introns are excised. RegTools[5] was used to extract splice junctions from aligned reads. LeafCutter[6] was subsequently employed to cluster overlapping intron excision events and quantify their relative excision ratios.

**Alternative transcription start site (TSS) and Alternative polyadenylation (polyA) usage**: To capture differential selection of transcription start or polyadenylation sites, which influences 5'UTR/3'UTR composition and post-transcriptional regulation, event-based transcript annotations were constructed using txrevise[7]. Subsequently, relative usage of these alternative sites was calculated via kallisto.

For each transcriptomic modality, a two-stage quantile normalization procedure was applied to generate standardized RNA trait tables. Let $X_{ij}$ denote the value of RNA trait $i$ ($i = 1, \ldots, p$) in sample $j$ ($j = 1, \ldots, n$). In the first step, trait values were quantile normalized across samples to standardize distributional effects among samples. For each sample $j$, trait values were sorted to obtain the ordered statistics $X_{(1)j} \leq X_{(2)j} \leq \cdots \leq X_{(p)j}$, and for each rank $k$, the average value across samples was calculated as $\bar{X}_{(k)} = \frac{1}{n}\sum_{j=1}^{n} X_{(k)j}$. Each trait value was then replaced by the average value corresponding to its rank position, such that

$$X'_{ij} = \bar{X}_{(k)},$$

where $k$ denotes the rank of $X_{ij}$ within sample $j$. In the second step, each transcriptomic trait was transformed using a rank-based inverse normal transformation across samples. For RNA trait $i$, values from the first step were ranked across samples to obtain $R_{ij}$ =rank ($X'_{ij}$), with ties preserved by assigning averaged ranks. Ranks were converted to $U_{ij} = \frac{R_{ij}}{n+1}$, which approximates the empirical cumulative distribution function (ECDF) of $X'_{ij}$ (i.e. $U_{ij} \approx \widehat{F}_i(X'_{ij})$). Finally, values were mapped to the standard normal distribution as

$$Z_{ij} = \Phi^{-1}(U_{ij}),$$

where $\Phi^{-1}$ denotes the inverse cumulative distribution function of the standard normal distribution, yielding approximately Gaussian-distributed RNA traits.

#### Conditionally independent cis-xQTL mapping by tensorQTL

Conditionally independent cis-xQTL mapping was executed using tensorQTL[8] via the Pheast pipeline for both separate- and cross-modality analyses. For separate-modality analyses, cis-xQTL mapping was performed independently for each of the six transcriptomic modalities using the normalized RNA trait tables as input. Covariates included the first 5 RNA trait principal components (PCs), the first 5 genotype

PCs, sex, education, age of death, study type (ROS or MAP), RNA Integrity Number (RIN), and batch effects. For modalities containing multiple molecular traits per gene (isoform ratios, splice junction usage, alternative TSS usage, and alternative polyA usage), all traits annotated to the same gene were analyzed as a single group so that cis-xQTLs were conditionally independent across RNA traits within each gene. For cross-modality analyses, normalized RNA trait tables from all six modalities were concatenated into a single trait table, and all traits corresponding to the same gene were assigned to the same group regardless of modality. A new set of RNA trait PCs was computed from the combined table and used as covariates together with the same covariates used for separate-modality analysis. Cis-xQTL mapping was then performed on this combined RNA trait table, jointly analyzing all traits assigned to each gene group so that cis-xQTLs were conditionally independent across all six transcriptomic modalities within each gene.

Initial standard QTL mapping was first conducted in cis mode by regressing each quantitative trait $y$ on variant genotype $X$ and the covariates matrix $C$. To identify the top associated variant for each trait group, permutation testing was performed using a beta-approximation to estimate empirical $p$-values. Storey's $q$-value was subsequently obtained from these empirical $p$-values to control the false discovery rate (FDR) at the gene-level ($q < 0.05$). Trait-group mapping files generated by Pheast were supplied to tensorQTL so that all molecular traits from the same gene (e.g. multiple exon junctions in splicing modality) were processed together as a single group during the mapping.

Next, cis_independent mode filtered for significant trait groups passing the gene-level FDR threshold ($q < 0.05$) and applied a stepwise regression procedure to each group. During forward selection, the algorithm identified the primary variant, which is the most significant variant within the defined cis-window. It then iteratively tested the remaining variants while conditioning on previously selected variants, continuing until no additional variants passed the significance threshold. Subsequently, a backward elimination was performed by re-testing each selected variant while conditioning on all other mapped independent signals in the model, removing any variants that lost statistical significance. All testing was based on empirical $p$-values estimated via permutation beta approximation. The variants retained after stepwise selection formed a single, ranked list of conditionally independent cis-xQTLs per gene, giving one or multiple independent variants within the same gene locus. The rank of each conditionally independent cis-xQTL was determined by its selection order during the stepwise selection process, with rank 1 indicating the primary (most significant) signal, and subsequent ranks representing secondary, independent signals with sequentially weaker effects.

#### Two-stage xTWAS analysis framework

Stage I of standard two-stage TWAS trains an imputation model for genetically regulated molecular traits using individual-level transcriptomic and genotype data from a reference dataset. The relationship between each transcriptional trait and genotypes is modelled using the following multiple linear regression model Eq. (1):

$$T_g = X_g w + \epsilon_g, \epsilon_g \sim N(0, I), I = diag(1,1, \dots ,1), \quad (1)$$

where $T_g$ denotes the transcriptional trait of a given gene $g$, $X_g$ denotes a $n \times m$ genotype matrix containing m cis-SNPs measured across $n$ individuals, $w$ represents a vector of genetic effect sizes (i.e., xQTL effect sizes in a broad sense), and $\epsilon_g$ is the error term. We considered only cis-SNPs located within 500kb around the transcription starting sites of the corresponding gene as a genotype predictor $X_g$. Prior to stage I, confounding covariates including 5 RNA trait PCs, 5 genotype PCs, sex, education, age of death, study type (ROS/MAP), RNA Integrity Number (RIN), and batch effects were regressed out from the original normalized molecular quantitative traits such that $T_g$ represents the regression residuals from that model. For each molecular trait, the genetic weight vector $\hat{w}$ was estimated using the best performing model, defined as the model achieving the highest cross-validation $R^2$ among the best linear unbiased predictor (BLUP), elastic net, lasso, and top single-variant (Top1) models implemented in FUSION[9].

In stage II of TWAS, given the estimated genetic weight vector $\hat{w}$ from stage I and independent individual-level GWAS data with genotype data $X_{new}$, the genetically regulated molecular trait for $X_{new}$ is imputed by $X_{new}\widehat{w}$. Then the association between genetically imputed trait $X_{new}\widehat{w}$ and the phenotype $Y$ is tested based on a generalized linear regression model as follows:

$$f(E[Y|X_{new}]) = \alpha(X_{new}\widehat{w}\,);\ H_0\colon \alpha = 0, H_a\colon \alpha \neq 0 \tag{2}$$

This gene-based association test can be equivalently performed with summary-level GWAS data by using the burden $Z$-score test statistic given by $\tilde{Z} = \frac{Z_{GWAS}\widehat{w}}{\sqrt{\widehat{w}'V\widehat{w}}}$, which has a standard normal distribution under the null hypothesis. Here, $Z_{GWAS}$ denotes a vector of single variant test $Z$-scores from GWAS summary data for all test SNPs in both FUSION weight panel and the GWAS dataset, and V denotes the genotype correlation matrix (i.e. linkage disequilibrium, LD) estimated from reference genotype data of the same ancestry.

#### ACAT for modalities that produce multiple transcriptomic

For transcriptomic modality generating multiple molecular traits per gene (e.g., alternative splicing, isoform ratios, alternative TSS usage, and alternative polyA usage), we applied Aggregated Cauchy Association Test[10] (ACAT) method to obtain a gene-level omnibus test p-value by aggregating trait-level TWAS p-values. The ACAT method first transforms p-values ($p_m$, $m$=1,...,$M$) obtained for all $M$ traits within a gene into variables following a standard Cauchy distribution, and then derives a test statistic by taking their average as follows:

$$T_{ACAT,g} = \frac{1}{M}\sum_{m=1}^{M} tan\{(0.5 - p_m)\pi\}, \pi \approx 3.14 \tag{3}$$

Under the same null hypothesis that the genetically regulated molecular traits within gene $g$ is not associated with the interested phenotype, ACAT test statistic can be approximated by a standard Cauchy distribution, and the corresponding omnibus ACAT p-value can be approximated as:

$$p_{ACAT,g} \approx \frac{1}{2} - \frac{\{\arctan(T_{Acat,g})\}}{\pi}, \pi \approx 3.14 \tag{4}$$

Previous studies[11,12] have demonstrated that ACAT provides improved statistical power while maintaining well-calibrated type I error rates.

#### Two-stage version of GIFT

To define the test regions for fine-mapping, we first extended the TSS of each significant TWAS risk gene by ±1Mb. Overlapping regions among adjacent TWAS risk genes were then merged into a single genomic region. Within each identified test region, we executed the two-stage version of GIFT[13] tool (*GIFT_two_stage_summ*), which enables the use of pre-computed xQTL weights and improve computational efficiency.

As described previously, the marginal TWAS Z-score for the $i$-th gene (or transcriptomic trait) is defined as $Z_{TWAS,i} = \frac{\widehat{\beta}_i}{se(\widehat{\beta}_i)} = \frac{z_{GWAS,i}w_i}{\sqrt{w_i^T V_i w_i}}$ , where $\hat{\beta}_i$ represents the marginal association of the $i$-th gene's predicted molecular traits with phenotype, $w_i$ represents the predictive cis-xQTL weights, $Z_{GWAS,i}$ represents the GWAS Z-scores for the corresponding cis-SNPs, and $V_i$ denotes the genotype correlation matrix among cis-SNPs. In the pre-process step of two-stage version of GIFT, these marginal TWAS Z-scores were concatenated into a regional vector, $Z_{TWAS} = (Z_{TWAS,1}, \ldots, Z_{TWAS,k})^T$, where k is the number of molecular traits in the locus. Concurrently, GWAS effect sizes and their corresponding standard errors were extracted for the cis-SNPs across all genes in the region.

Regional LD matrices (V) containing cis-SNPs within the locus were generated using PLINK2[14](v2.00a4.3), applying quality control filters to retain variants with a minor allele frequency (MAF) > 0.01 and a genotype missingness rate < 0.1 to avoid computational instability. Since the use of an out-of-sample LD reference panel can introduce noise, LD matrix was regularized using susieR[15] to $V_{reg} = (1 - s)V + sI$, where I is the identity matrix and s denotes the shrinkage parameter estimated via maximum likelihood based on the GWAS summary data. Subsequently, the predicted variance-covariance matrix of the genetically regulated molecular traits ($\hat{R}$) for all k genes in the test region is derived as $\hat{R} = W^T V_{reg} W$, where $W = w_1, \ldots, w_k)^T$ represents the concatenated cis-xQTL weights across k traits in the region.

Then, the joint conditional effect sizes ($\hat{\beta}_{cond}$ ) for all genes were subsequently estimated as:

$$\hat{\beta}_{cond} = \hat{R}^{-1}\hat{\beta} = (W^T V_{reg} W)^{-1}(Z_{GWAS} W),$$

where $\hat{\beta}$ is a $k$-vector of marginal gene effect estimates from marginal TWAS analysis. By adjusting the marginal associations by the inverse of the covariance structure, the model explicitly conditions on the correlation structure among molecular traits and cis-SNP LD across multiple nearby genes. This formulation yields a final conditional Z-score vector that is equivalent to the naïve adjustment approach, given by $Z_{TWAS_{cond}} = \hat{R}^{-1/2} Z_{TWAS}$. Corresponding two-tailed p-values were subsequently derived from these final conditional Z-scores to identify putatively causal genes.

### Supplemental References


1. Mölder, F., Jablonski, K.P., Letcher, B., Hall, M.B., van Dyken, P.C., Tomkins-Tinch, C.H., Sochat, V., Forster, J., Vieira, F.G., Meesters, C., et al. (2021). Sustainable data analysis with Snakemake. F1000Res *10*, 33. https://doi.org/10.12688/f1000research.29032.3.

2. Munro, D., Ehsan, N., Esmaeili-Fard, S.M., Gusev, A., Palmer, A.A., and Mohammadi, P. (2024). Multimodal analysis of RNA sequencing data powers discovery of complex trait genetics. Nat Commun *15*, 10387. https://doi.org/10.1038/s41467-024-54840-8.

3. Bray, N.L., Pimentel, H., Melsted, P., and Pachter, L. (2016). Near-optimal probabilistic RNA-seq quantification. Nat Biotechnol *34*, 525–527. https://doi.org/10.1038/nbt.3519.

4. Liao, Y., Smyth, G.K., and Shi, W. (2013). The Subread aligner: fast, accurate and scalable read mapping by seed-and-vote. Nucleic Acids Res *41*, e108. https://doi.org/10.1093/nar/gkt214.

5. Cotto, K.C., Feng, Y.-Y., Ramu, A., Richters, M., Freshour, S.L., Skidmore, Z.L., Xia, H., McMichael, J.F., Kunisaki, J., Campbell, K.M., et al. (2023). Integrated analysis of genomic and transcriptomic data for the discovery of splice-associated variants in cancer. Nat Commun *14*, 1589. https://doi.org/10.1038/s41467-023-37266-6.

6. Li, Y.I., Knowles, D.A., Humphrey, J., Barbeira, A.N., Dickinson, S.P., Im, H.K., and Pritchard, J.K. (2018). Annotation-free quantification of RNA splicing using LeafCutter. Nat Genet *50*, 151–158. https://doi.org/10.1038/s41588-017-0004-9.

7. Alasoo, K., Rodrigues, J., Danesh, J., Freitag, D.F., Paul, D.S., and Gaffney, D.J. (2019). Genetic effects on promoter usage are highly context-specific and contribute to complex traits. Elife *8*, e41673. https://doi.org/10.7554/eLife.41673.

8. Taylor-Weiner, A., Aguet, F., Haradhvala, N.J., Gosai, S., Anand, S., Kim, J., Ardlie, K., Van Allen, E.M., and Getz, G. (2019). Scaling computational genomics to millions of individuals with GPUs. Genome Biol *20*, 228. https://doi.org/10.1186/s13059-019-1836-7.

9. Gusev, A., Ko, A., Shi, H., Bhatia, G., Chung, W., Penninx, B.W.J.H., Jansen, R., de Geus, E.J.C., Boomsma, D.I., Wright, F.A., et al. (2016). Integrative approaches for large-scale transcriptome-wide association studies. Nat Genet *48*, 245–252. https://doi.org/10.1038/ng.3506.

10. Liu, Y., Chen, S., Li, Z., Morrison, A.C., Boerwinkle, E., and Lin, X. (2019). ACAT: A Fast and Powerful p Value Combination Method for Rare-Variant Analysis in Sequencing Studies. The American Journal of Human Genetics *104*, 410–421. https://doi.org/10.1016/j.ajhg.2019.01.002.

11. Guo, S., and Yang, J. (2024). Bayesian genome-wide TWAS with reference transcriptomic data of brain and blood tissues identified 141 risk genes for Alzheimer’s disease dementia. Alz Res Therapy *16*, 120. https://doi.org/10.1186/s13195-024-01488-7.

12. Hu, T., Parrish, R.L., Dai, Q., Buchman, A.S., Tasaki, S., Bennett, D.A., Seyfried, N.T., Epstein, M.P., and Yang, J. (2024). Omnibus proteome-wide association study identifies 43 risk genes for Alzheimer disease dementia. The American Journal of Human Genetics *111*, 1848–1863. https://doi.org/10.1016/j.ajhg.2024.07.001.

13. Liu, L., Yan, R., Guo, P., Ji, J., Gong, W., Xue, F., Yuan, Z., and Zhou, X. (2024). Conditional transcriptome-wide association study for fine-mapping candidate causal genes. Nat Genet *56*, 348–356. https://doi.org/10.1038/s41588-023-01645-y.

14. Chang, C.C., Chow, C.C., Tellier, L.C., Vattikuti, S., Purcell, S.M., and Lee, J.J. (2015). Second-generation PLINK: rising to the challenge of larger and richer datasets. Gigascience *4*, s13742-015-0047–0048. https://doi.org/10.1186/s13742-015-0047-8.

15. Wang, G., Sarkar, A., Carbonetto, P., and Stephens, M. (2020). A simple new approach to variable selection in regression, with application to genetic fine mapping. Journal of the Royal Statistical Society: Series B (Statistical Methodology) *82*, 1273–1300. https://doi.org/10.1111/rssb.12388.
